# Natalizumab Treatment Continuum in RRMS: A Systematic Review and Meta-Analysis of Cessation Risks and Dosing Strategies

**DOI:** 10.1101/2025.10.20.25338391

**Authors:** Vihaan Sahu, Mohith Balakrishnan, Dhaneesh Rucher Jetty

**Affiliations:** Georgian National University SEU, Tbilisi, Georgia

**Keywords:** Natalizumab, multiple sclerosis, relapsing-remitting, extended interval dosage, treatment discontinuation, meta-analysis, progressive multifocal leukoencephalopathy

## Abstract

**Background:** Natalizumab is a high-efficacy therapy for relapsing-remitting multiple sclerosis (RRMS). While extended interval dosing (EID) may reduce progressive multifocal leukoencephalopathy (PML) risk, optimizing post-cessation outcomes remains critical for long-term management.

**Objectives:** To compare the efficacy and safety of natalizumab standard interval dosing (SID) versus EID and to quantify relapse and disability risk after cessation in adults with RRMS.

**Methods:** We conducted a systematic review and meta-analysis (PROSPERO: CRD420251103014) following PRISMA guidelines. MEDLINE, Cochrane Library, ClinicalTrials.gov, and other sources (2015–2025) were searched for randomized controlled trials and observational studies. Primary outcomes were annualized relapse rate (ARR) and disability progression; secondary outcomes included PML incidence and post-cessation relapse. Risk of bias was assessed using Cochrane RoB 2.0 and ROBINS-I. Data were synthesized narratively or via meta-analysis, with evidence certainty graded using GRADE.

**Results:** Twenty-eight studies (n=47,987) were included. Pooled relapse risk within 12 months of cessation was 41.7% (95% CI 30.6 to 53.6%) based on 4 studies (n=861); sensitivity analysis excluding the outlier (Weinstock-Guttman 2016) showed 36.8%. Narrative synthesis of ARR (9 studies) showed no consistent SID versus EID difference (median difference 0.00). Meta-analysis of PML risk (3 studies) found no significant difference (RR 0.70, 95% CI 0.20–2.54). Disability outcomes were too heterogeneous for meta-analysis. Evidence certainty was low for most outcomes.

**Conclusion:** Natalizumab cessation carries a high relapse risk. No consistent efficacy difference between EID and SID was identified, although formal equivalence or non-inferiority was not tested; relapse rates underscore the importance of planning discontinuation.

## 1. Introduction

### 1.1 Background and Significance

Multiple sclerosis (MS) is a chronic immune-mediated disease causing neurological disability. Globally affecting 2.8 million people, RRMS accounts for 85-90% of cases and typically presents in early adulthood (1–3). Effective disease-modifying therapy is crucial for managing relapses and disability progression. Hospitalization and large medical expenses are common among patients with severe RRMS, emphasizing the need for effective treatment options (4, 5).

Natalizumab, a humanized monoclonal antibody targeting α4-integrin, inhibits lymphocytes from crossing the blood-brain barrier, reducing CNS inflammation and relapse activity (6, 7). Since its FDA approval in 2004, natalizumab has demonstrated significant decreases in relapse frequency and MRI lesion burden in pivotal trials (8, 3, 2). Following reports of PML in 2005, risk management strategies such as patient screening and JC virus surveillance were implemented (9, 10). Despite these developments, questions about optimal dosing regimens and safe withdrawal continue to arise. While standard four-week dosing maintains α4-integrin receptor saturation above 80%, extended interval dosing (EID, 5 to 8 weeks) may reduce PML risk while maintaining efficacy (5). Discontinuation carries a 3-6-month risk of disease reactivation, highlighting the need for evidence-based management during this period. Clinical decisions regarding dosing intervals are often linked to planning for eventual withdrawal, necessitating a comprehensive review of the treatment continuum.

### 1.2 Current Evidence and Gaps

Phase 2 and pivotal studies consistently demonstrated lower relapse rates and MRI lesion activity with natalizumab compared to controls (8, 3, 2, 11). Early meta-analyses confirmed efficacy but were restricted by short follow-up periods and a lack of long-term safety data. Nikfar et al. (12) (2010) analyzed four studies (n = 1,407) and reported a 50% reduction in relapse risk at the approved dose (RR 0.5, 95% CI 0.42 to 0.61), while the pooled estimate across all doses was 0.7 (0.42 to 1.17); the applicability to modern extended dosing regimens was not specified. Subsequent observational studies, such as Salhofer-Polanyi et al. (13) (2014), indicated the risk of disease reactivation following cessation; however, variability in study designs and outcome definitions has limited the ability to draw definitive conclusions. While Rabea et al. (14) (2024) provided valuable insights into natalizumab dosing strategies, their review did not address post-cessation outcomes. This is a notable gap because clinical decisions regarding dosing intervals are often linked to withdrawal planning.

Current EMA guidelines recommend that treatment plans include protocols for both ongoing dosing and eventual discontinuation (54). Therefore, an integrated analysis of both dosing and cessation outcomes is relevant for clinical decision-making. The present review expands upon previous work by integrating analysis of dosing strategies with a comprehensive synthesis of post-cessation relapse risks, utilizing updated data through 2025.

### 1.3 Rationale for this Review

While natalizumab’s efficacy is established, integrating evidence on dosing strategies with post-discontinuation outcomes provides a more comprehensive view of the treatment continuum. Real-world evidence suggests a risk of rebound, but quantifying this risk across diverse study designs is necessary. We provide a systematic analysis of post-cessation outcomes alongside updated evidence on dosing strategies to support clinical decision-making regarding the full duration of therapy.

### 1.4 Objectives

This systematic review and meta-analysis aims to provide comprehensive evidence on natalizumab treatment in adults with RRMS, with a primary focus on post-cessation outcomes and updated analysis of dosing strategies. Using a PICO framework, the objectives are:

- To quantify the absolute risk, timing, and severity of relapse and disability progression following treatment cessation (primary focus).
- To compare the efficacy of standard-interval dosing (SID) versus extended-interval dosing (EID) on annualized relapse rate (ARR), disability progression, and MRI outcomes.
- To evaluate safety profiles, particularly the incidence of PML.
- To synthesize real-world and trial evidence from 2015 onward.
- To support clinical decision-making by offering evidence on dose optimization and cessation procedures.

## 2. Methods

### 2.1 Protocol and Registration

This systematic review was conducted according to a predefined protocol registered on PROSPERO (CRD420251103014). The protocol specified the search strategy, eligibility criteria, outcomes, risk-of-bias assessment methods, and synthesis approach. The review is reported in accordance with the PRISMA 2020 statement (15), which updated the original 2009 statement (18).

### 2.2 Eligibility Criteria

Studies were selected based on the PICO framework:

- Population: Adults (≥18 years) with relapsing-remitting multiple sclerosis (RRMS) treated with natalizumab monotherapy.
- Intervention: 1) Standard-interval dosing (SID) of natalizumab (every 4 weeks), or 2) Cessation of natalizumab therapy.
- Comparator: 1) Extended-interval dosing (EID; infusion intervals typically >4 weeks, up to 8 weeks), or 2) For cessation studies, either continuation of natalizumab or no active comparator (natural history).
- Outcomes: The primary outcomes were the annualized relapse rate (ARR) and disability progression (confirmed EDSS deterioration). Secondary outcomes included No Evidence of Disease Activity (NEDA-3), relapse risk after cessation, MRI activity, and safety outcomes, specifically the incidence of progressive multifocal leukoencephalopathy (PML).
- Study Designs: Randomized controlled trials (RCTs), prospective and retrospective cohort studies, and case series (for cessation results) were all included. Studies were omitted if they were reviews, addressed pediatric populations, coupled natalizumab with other disease-modifying treatments, or did not provide relevant outcomes. Because some included cohorts enrolled mixed populations, studies were eligible when patients with RRMS constituted the majority of the cohort or when RRMS-specific outcomes were reported separately; this applied to three studies (39, 43, 52) and is acknowledged in Section 4.3. To reflect current practice, the publishing date range began on January 1, 2015. While the protocol specified studies from 2015 onward, one study (16) was first published online in November 2014 but assigned to a 2015 journal issue; given its direct relevance, it was retained. This represents a minor deviation from the protocol. The protocol excluded conference abstracts without full data; two included studies (28, 47) are congress abstracts with extractable data, retained for their direct relevance. Only English-language publications were included. For the comparator definition, EID was treated as a heterogeneous exposure (mean or planned interval of 5 to 8 weeks), because an interval of five weeks is not equivalent to an interval of eight weeks; study level definitions are reported in Tables 1 and 2A, and the influence of interval length was examined descriptively in Section 3.4.1.

**Table 1.** Characteristics of Included Studies.

| Study (Author, Year) | Country | Study Design | Sample Size (Total N) | Comparison Groups | Funding Source / Conflict of Interest | Notes |
|---|---|---|---|---|---|---|
| Al-Mahdawi et al., 2023 (28) | Iraq | Observational Retrospective Cohort | 150 | SID (Q4W), EID (definition NR) | NR | Multicenter, aggregated data only (contributed to narrative context) |
| Bernardes et al., 2024 (29) | Portugal | Retrospective observational cohort | 76 | SID (Q4W) vs EID (Q6W) | No specific grant; COI disclosed | Single-center, within-patient comparison |
| Butzkueven et al., 2021 (TOP) (30) | Multinational | Prospective Observational Registry | 472 (236 pairs) | SID (Q4W), EID (Q6W) | Supported by Biogen; multiple author COIs | Multicenter registry, propensity score-matched |
| Clerico et al., 2020 (31) | Italy | Retrospective Cohort | 360 | SID (<5 weeks), EID (≥5 weeks) | NR | Multicenter (14 centers) |
| Coerver et al., 2021 (32) | Netherlands | Retrospective Observational Cohort | 147 | Cessation | ZonMW grant, Stichting MS Research grant | Monocentric, outcomes after cessation |
| De Mercanti et al., 2021 (33) | Italy | Retrospective cohort | 316 | SID (<5w) vs EID (≥5w) | Not reported | Multicenter (14 centers) |
| Foley et al., 2022 (NOVA) (34) | Multinational (11 countries) | RCT (open-label, rater-blinded) | 499 | SID (Q4W) vs EID (Q6W) | Biogen | Multicenter (89 centers), phase 3b trial |
| Fuentes-Rumí et al., 2020 (35) | Spain | Retrospective observational | 50 | Cessation | No specific grant; COI disclosed | Single-center, IVMP washout strategies |
| Gardinetti et al., 2017 (36) | Italy | Retrospective Cohort | 30 | Cessation | All authors have nothing to declare | Multicenter (2 centers), outcomes after cessation |
| González-Suarez et al., 2017 (37) | Spain | Case series | 4 | Cessation → switch to FTY | "No conflict of interest" | Multicenter (3 hospitals), rebound cases |
| Haas et al., 2017 (38) | Germany | Prospective cohort | 57 | SID (Q4W) vs Cessation (Wash-out) | German Ministry for Education and Research; "none declared" | Single-center, immunology focus |
| Jakimovski et al., 2023 (39) | USA (NY) | Retrospective cohort | 242 | Continue SID (Q4W) vs Discontinue (Cessation) | Biogen; Multiple authors disclosed industry funding | Consortium data, includes SPMS/PPMS patients |
| Jeantin et al., 2023 (40) | France | Retrospective Self-Controlled Cohort | 57 | SID (Q4W), EID (Q6W) | No funding; some author COIs | Monocentric, within-patient comparison |
| Kaufman et al., 2015 (RESTORE) (41) | Multinational | RCT | 175 | Continue Natalizumab (Q4W), Placebo (Cessation) | Funded by Biogen Idec Inc. | Multicenter, partially placebo-controlled |
| Kister et al., 2018 (42) | Multinational (20 centers) | Retrospective registry cohort | 4842 | Cessation (vs other DMTs) | MSBase Foundation; "No targeted funding" | MSBase registry, natalizumab cessation arm n=308 |
| Mustonen et al., 2020 (43) | Finland | Retrospective cohort | 89 (85 RRMS) | Cessation | Hospital grants; personal grants; honoraria/consultancies | Multicenter (4 hospitals), RRMS n=85 for PICO |
| Papeix et al., 2016 (44) | France | Prospective observational registry | 715 | Cessation | No targeted funding; COI disclosed | Nationwide multicenter (TYSEDMUS registry) |
| Pelle et al., 2023 (45) | France | Retrospective cohort | 303 | SID (Q4W) vs EID (5-6w) | Caen University Hospital; additional funding from Biogen | Multicenter (5 centers) |
| Riancho et al., 2021 (46) | Spain | Retrospective Cohort | 39 | SID (Q4W), EID (Q8W) | Supported by IDIVAL; various author COIs | Monocentric, within-patient comparison (sequential arms) |
| Ruggieri et al., 2019 (47) | Italy | Retrospective Cohort | 775 | SID (≤35 days), EID (>35 and ≤56 days) | NR | Multicenter, includes a mixed-dose arm |
| Ryerson et al., 2016 (48) | USA | Registry Analysis | 1998 | SID (Q4W ±2d), EID (4w3d-8w5d) | No grant or industry support | Multicenter registry |
| Ryerson et al., 2019 (27) | USA | Retrospective Cohort (TOUCH) | 35,521 | SID (various def.), EID (various def.) | Funded by Biogen | TOUCH registry, analysis of PML risk only |
| Ruggieri et al., 2024 (49) | Italy | Retrospective Cohort | 745 | SID (Q4W), EID (>5-7 weeks), EID (>7-8 weeks) | Multiple author COIs with pharma companies | Multicenter (8 centers), SID n=627, EID n=118 |
| Toorop et al., 2024 (50) | Netherlands | Retrospective Cohort | 82 | SID (Q4W), EID (≥5 weeks) | Dutch MS Research Foundation, Brain Foundation Netherlands, Innovation Fund Healthcare insurers | Monocentric |
| van Kempen et al., 2020 (51) | Netherlands | Prospective single-arm trial | 61 | EID (Personalized) | Brain Foundation Netherlands; Various COI disclosures | Multicenter, no SID comparator arm |
| Vidal-Jordana et al., 2015 (16) | Spain | Prospective observational cohort | 47 | Cessation | No specific grant | Single-center, outcomes after cessation |
| Weinstock-Guttman et al., 2016 (52) | USA | RCT (single-blinded MRI) | 50 | Cessation: Tapered (EID-like) vs Immediate | Biogen Idec (Investigator-initiated) | Single-center, phase IV, includes relapsing-SPMS |
| Yamout et al., 2018 (53) | Lebanon, Iran | Retrospective Review of Registry Cohorts | 85 | SID (Q4W), EID (≥5 weeks) | None | Multicenter (2 centers), within-patient comparison |

*Footnote: SID = standard-interval dosing; EID = extended-interval dosing; RRMS = relapsing-remitting multiple sclerosis; SPMS = secondary progressive multiple sclerosis; PPMS = primary progressive multiple sclerosis; NR = not reported.*

**Table 2A:** ARR Comparison: Study Characteristics and Reported Outcomes.

| Study | EID Definition | Reported ARR (Quote/Direct Summary) | Notes |
|---|---|---|---|
| Bernardes 2024 (29) | 6 weeks | ARR, median (IQR) 0 (0) [SID] vs 0 (0) [EID]; Relapses: 1 [SID] vs 4 [EID], p=0.083 | Within-patient comparison; PY not reported |
| Clerico 2020 (31) | ≥5 weeks | Adjusted ARR: 0.060 (95% CI 0.033–0.087) [SID] vs 0.039 (95% CI 0.017–0.063) [EID] | Adjusted mean; no raw counts/PY |
| De Mercanti 2021 (33) | ≥5 weeks | ARR 0.075 (SD 0.20) [SID] vs 0.089 (SD 0.22) [EID] | Pre-intervention period |
| Foley 2022 (34) | 6 weeks | Unadjusted ARR 0.0171 [SID] vs 0.0228 [EID]; Adjusted ARR: 0.00010 (95% CI 0.00004–0.00024) [SID] vs 0.00013 (95% CI 0.00006–0.00027) [EID] | Relapse counts not reported |
| Jeantin 2023 (40) | 6 weeks (42 ± 7 days) | ARR 0.0 [SID] vs 0.0 [EID] | Zero relapses; PY calculable |
| Pelle 2023 (45) | 5–6 weeks (≤9 infusions/12mo) | ARR 0.083 (SD 0.300) [SID] vs 0.041 (SD 0.231) [EID]; Patients with ≥1 relapse: 12/156 [SID] vs 5/147 [EID] | Only study with counts & PY suitable for a formal comparison |
| Riancho 2021 (46) | 8 weeks | ARR 0.025 (SD 0.026) [SID] vs 0.020 (SD 0.060) [EID]; p=0.72 | Mean ARR only |
| Ryerson 2016 (48) | 4.5–8.5 weeks | ARR 0.14 [SID] vs 0.09 [EID] | Mean ARR only |
| Yamout 2018 (53) | ≥5 weeks | ARR 0.0006 ± 0.003 [SID] vs 0.001 ± 0.006 [EID]; p=0.42 | Mean ARR and SD |

*Footnotes: SID = standard-interval dosing; EID = extended-interval dosing; ARR = annualized relapse rate; SD = standard deviation; PY = person-years; CI = confidence interval.*

**Table 2B:** Disability Progression and NEDA-3 (SID versus EID and After Cessation)

| Outcome | PICO Comparison | Study | Outcome Definition | Reported Results | Notes |
|---|---|---|---|---|---|
| EDSS Worsening | SID vs EID | Foley 2022 (34) | 24-week confirmed disability worsening | % free of progression: 92% [SID] vs 90% [EID] | Raw event counts not reported. |
|  | SID vs EID | Pelle 2023 (45) | Worsening of EDSS score (not confirmed) | 20 (13%) [SID] vs 14 (11%) [EID] | Definition is not confirmed disability progression. |
|  | After Cessation | Vidal-Jordana 2015 (16) | Increase of at least 2.0 points (or at least 1.0 point if the EDSS at withdrawal was 5.5 or higher), assessed 6 to 12 months after withdrawal | Eight patients (19%) presented significant clinical worsening | Single arm reporting only. |
| NEDA-3 | SID vs EID | Foley 2022 (34) | No relapses, no new MRI lesions, no 24-wk CDW | NEDA-3: 163/242 [SID] vs 173/247 [EID] | Only study reporting complete NEDA-3 data. |
|  | SID vs EID | Riancho et al., 2021 (46) | Not fully defined | Proportions for EID arm only (94%, 73%, 70%) | Incomplete reporting; no SID data. |

*Footnote: EDSS = Expanded Disability Status Scale; CDW = Confirmed Disability Worsening; NEDA-3 = No Evidence of Disease Activity (relapses, MRI activity, and CDW)*

**Table 2C:** PML Incidence in Patients on Standard-Interval Dosing (SID) vs. Extended-Interval Dosing (EID)

| STUDY (FIRST AUTHOR, YEAR) | DOSING GROUP | POPULATION AT RISK | PML EVENTS (N) | EXPOSURE DURATION | KEY NOTES |
|---|---|---|---|---|---|
| Studies Included in RR Meta-analysis |  |  |  |  |  |
| Butzkueven et al., 2021 (30) | SID | 210 (N) | 2 | Mean follow-up ~2 years | Safety population |
|  | EID | 236 (N) | 2 | Mean follow-up ~2 years | Safety population |
| Ryerson et al., 2019 (27) | SID | 4,801 (N)* | 19 | 49 to 60 infusions | Adjusted JCV+ patients without prior immunosuppressant use |
|  | EID | 810 (N) | 1 | 49 to 60 infusions | Adjusted JCV+ patients without prior immunosuppressant use |
| Foley et al., 2022 (34) | SID | 242 (N) | 0 | 72 weeks (~1.4 years) | Clinical & MRI monitoring |
|  | EID | 247 (N) | 1 (asymptomatic) | 72 weeks (~1.4 years) | Asymptomatic case via MRI |
| Study with Person-Year Data |  |  |  |  |  |
| Ryerson et al., 2016 (48) | SID | 1052 PY** | 4 | JCV+ PY | JCV+ person-years |
|  | EID | 1090 PY** | 0 | JCV+ PY | JCV+ person-years |
| Studies with Zero PML Events |  |  |  |  |  |
| Clerico et al., 2020 (31) | SID | 216 (N) | 0 | Up to 2 years | Follow-up up to 2 years |
|  | EID | 144 (N) | 0 | Up to 2 years | Follow-up up to 2 years |
| Pelle et al., 2023 (45) | SID | 156 (N) | 0 | 12 months | 12 months follow-up |
| STUDY (FIRST AUTHOR, YEAR) | DOSING GROUP | POPULATION AT RISK | PML EVENTS (N) | EXPOSURE DURATION | KEY NOTES |
|  | EID | 147 (N) | 0 | 12 months | 12 months follow-up |
| Riancho et al., 2021 (46) | SID | 39 (N) | 0 | Mean 51.1 months | Mean follow-up |
|  | EID | 39 (N) | 0 | Mean 76.7 months | Mean follow-up |
| Toorop et al., 2024 (50) | SID | 50 (N) | NR | NR | PML not reported |
|  | EID | 32 (N) | NR | NR | PML not reported |
| Yamout et al., 2018 (53) | SID | 85 (N) | 0 | Not specified | Follow-up not specified |
|  | EID | 85 (N) | 0 | Not specified | Follow-up not specified |

Footnote: EID: Extended-Interval Dosing; JCV: JC virus; MRI: Magnetic Resonance Imaging; N: Number of patients; PML: Progressive Multifocal Leukoencephalopathy; PY: Person-Years; SID: Standard-Interval Dosing. \* Events and denominators refer to the conditional PML risk at infusions 49 to 60 (epoch 5) among anti-JC virus antibody positive patients without prior immunosuppressant use, reported by the study as 3.96 per 1,000 patients (SID) and 1.23 per 1,000 patients (EID); these counts were used for the pooled RR. \*\* Person-years of exposure.

**Table 2D:**
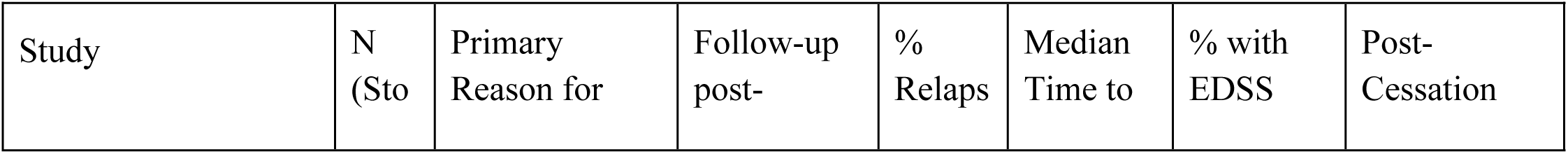
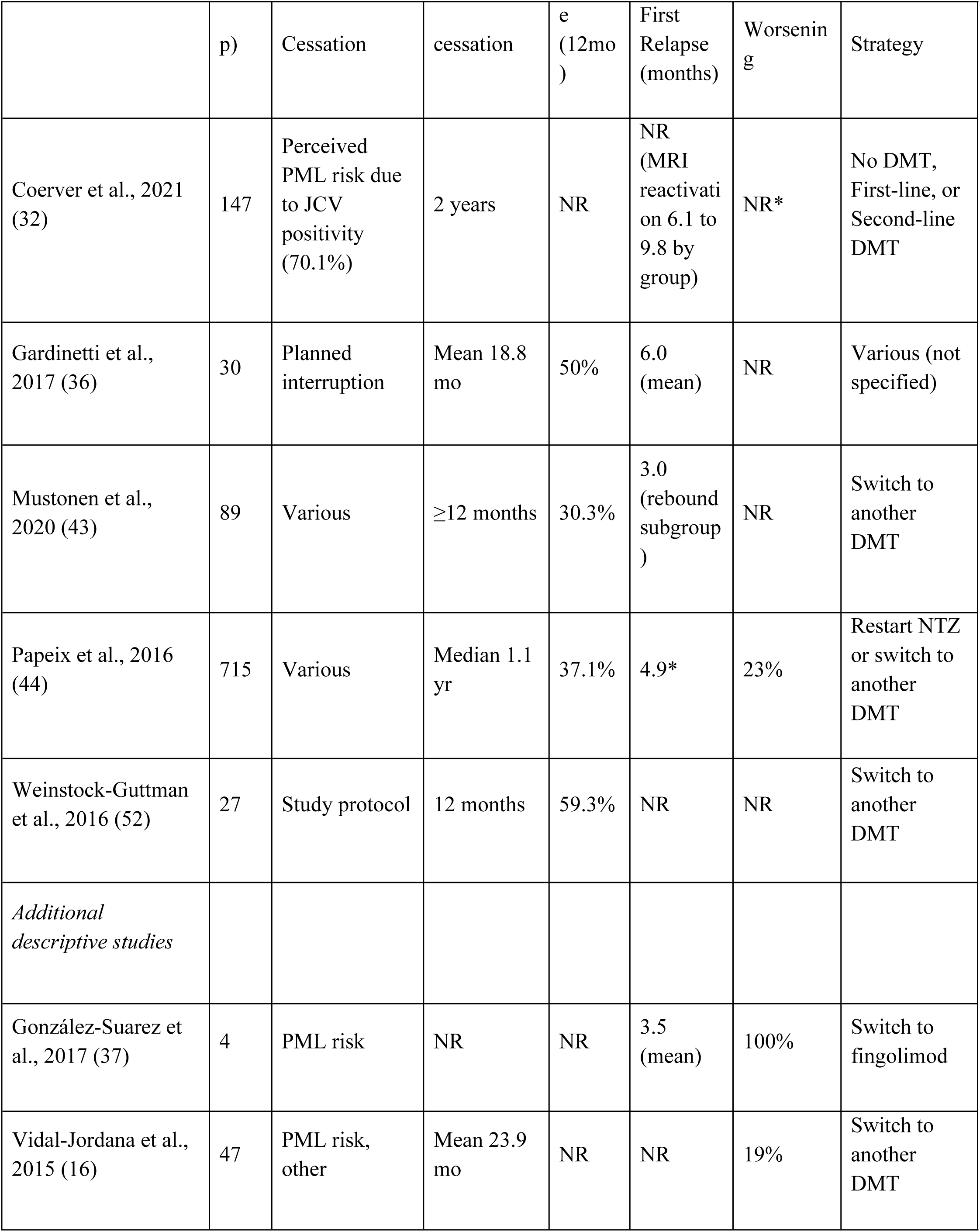
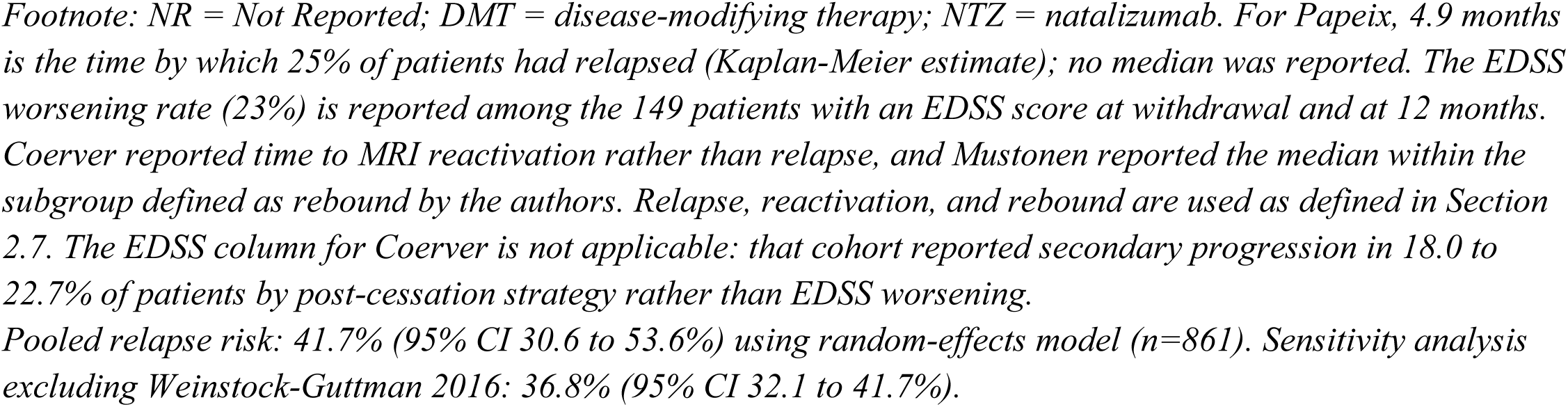
Outcomes After Natalizumab Cessation.

| Study | N (Sto | Primary Reason for | Follow-up post- | % Relaps | Median Time to | % with EDSS | Post-Cessation |
|---|---|---|---|---|---|---|---|
|  | p) | Cessation | cessation | e (12mo) | First Relapse (months) | Worsening | Strategy |
| Coerver et al., 2021 (32) | 147 | Perceived PML risk due to JCV positivity (70.1%) | 2 years | NR | NR (MRI reactivation on 6.1 to 9.8 by group) | NR* | No DMT, First-line, or Second-line DMT |
| Gardinetti et al., 2017 (36) | 30 | Planned interruption | Mean 18.8 mo | 50% | 6.0 (mean) | NR | Various (not specified) |
| Mustonen et al., 2020 (43) | 89 | Various | ≥12 months | 30.3% | 3.0 (rebound subgroup) | NR | Switch to another DMT |
| Papeix et al., 2016 (44) | 715 | Various | Median 1.1 yr | 37.1% | 4.9* | 23% | Restart NTZ or switch to another DMT |
| Weinstock-Guttman et al., 2016 (52) | 27 | Study protocol | 12 months | 59.3% | NR | NR | Switch to another DMT |
| *Additional descriptive studies* |  |  |  |  |  |  |  |
| González-Suarez et al., 2017 (37) | 4 | PML risk | NR | NR | 3.5 (mean) | 100% | Switch to fingolimod |
| Vidal-Jordana et al., 2015 (16) | 47 | PML risk, other | Mean 23.9 mo | NR | NR | 19% | Switch to another DMT |

*Footnote: NR = Not Reported; DMT = disease-modifying therapy; NTZ = natalizumab. For Papeix, 4.9 months is the time by which 25% of patients had relapsed (Kaplan-Meier estimate); no median was reported. The EDSS worsening rate (23%) is reported among the 149 patients with an EDSS score at withdrawal and at 12 months. Coerver reported time to MRI reactivation rather than relapse, and Mustonen reported the median within the subgroup defined as rebound by the authors. Relapse, reactivation, and rebound are used as defined in Section 2.7. The EDSS column for Coerver is not applicable: that cohort reported secondary progression in 18.0 to 22.7% of patients by post-cessation strategy rather than EDSS worsening.*
*Pooled relapse risk: 41.7% (95% CI 30.6 to 53.6%) using random-effects model (n=861). Sensitivity analysis excluding Weinstock-Guttman 2016: 36.8% (95% CI 32.1 to 41.7%).*

### 2.3 Information Sources

We searched MEDLINE (via PubMed), the Cochrane Library, ClinicalTrials.gov, Google Scholar, Sage Journals, Frontiers, eLife, BMJ, and DOAJ from 2015 to 2025, matching the sources summarized in the PRISMA flow diagram (Figure 1); additional studies were identified through reference screening and author contact.

**Figure 1:**
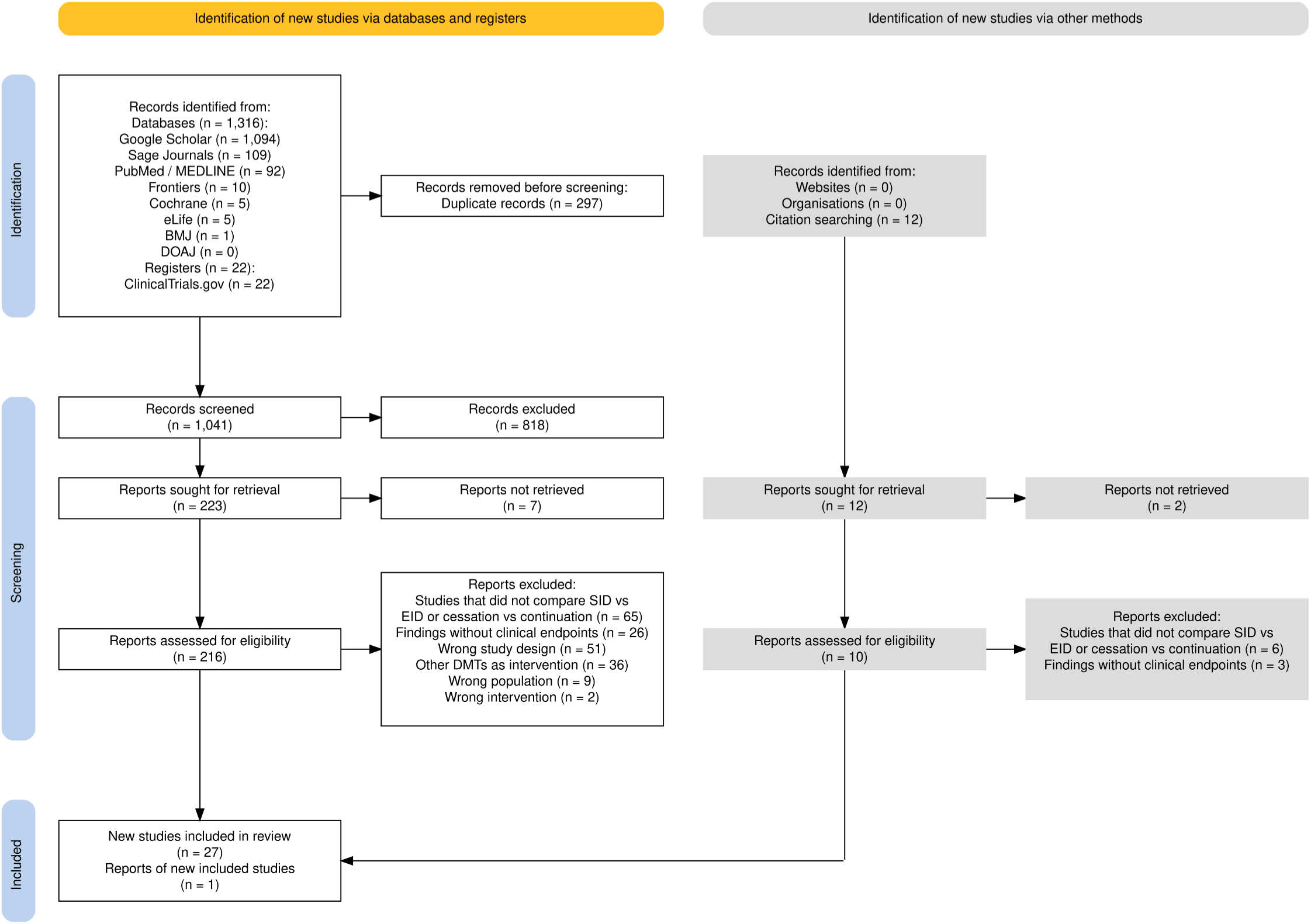
PRISMA flow chart

**Figure 2.**
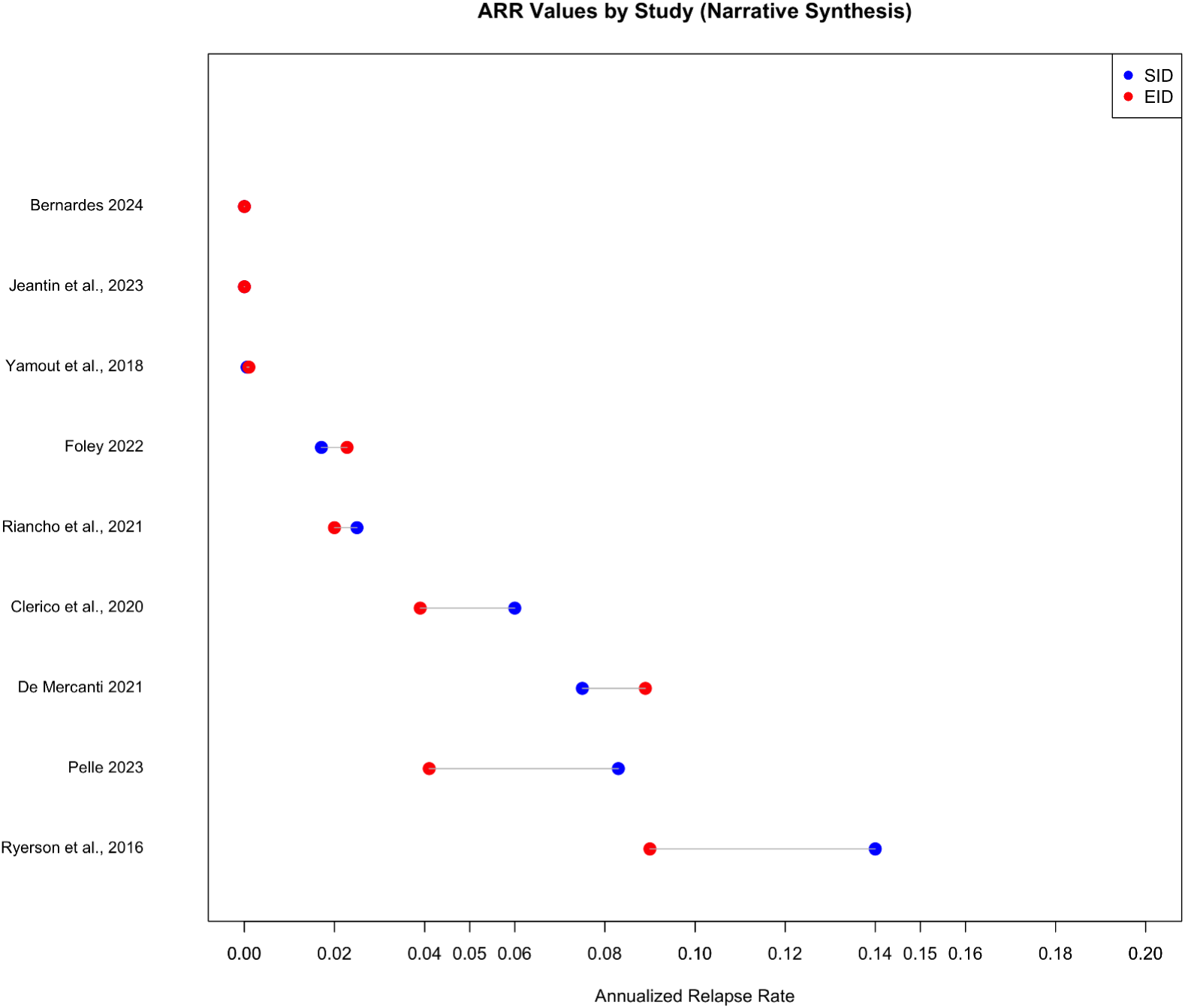
Annualized Relapse Rate (ARR) Values by Study (Narrative Synthesis).

**Figure 3.**
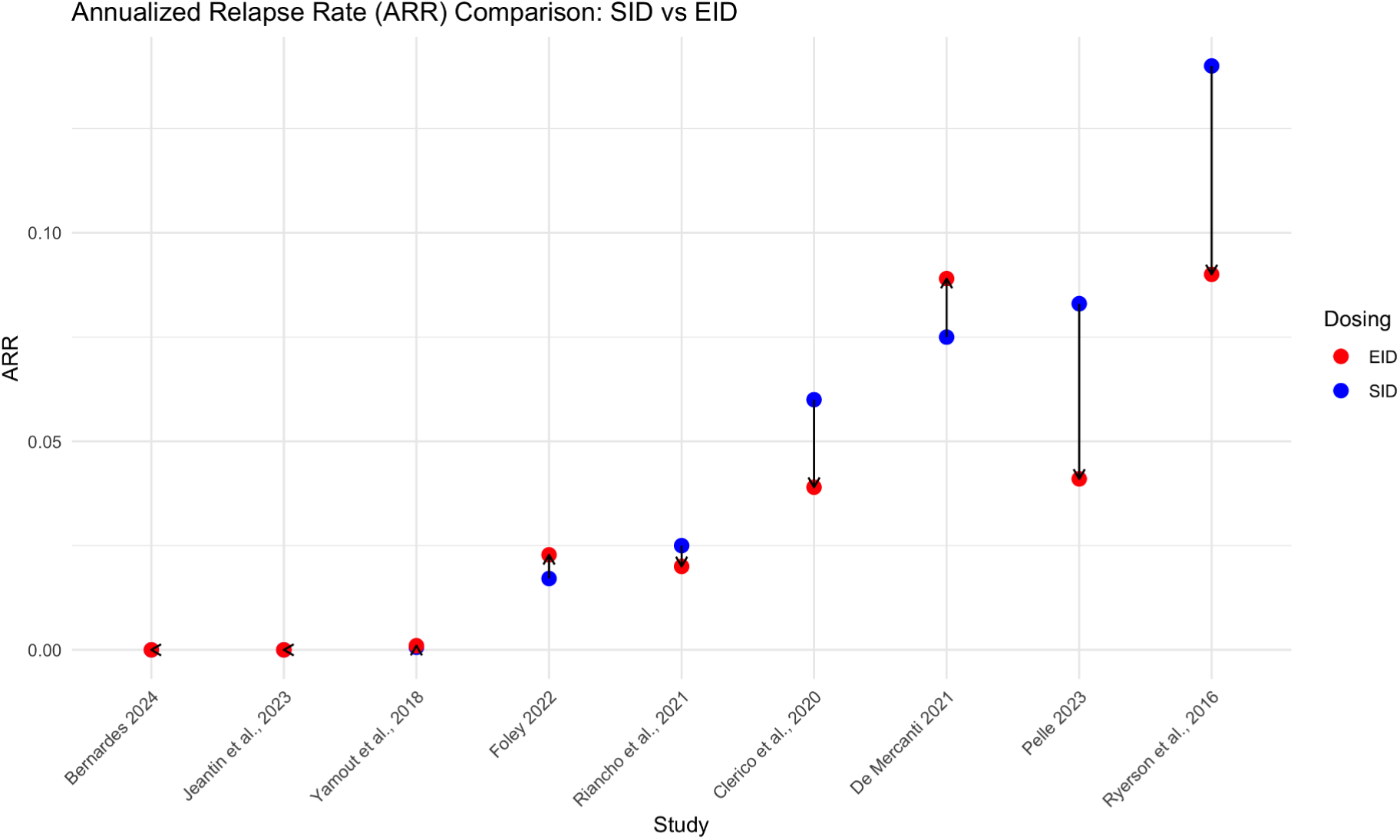
Annualized Relapse Rate (ARR) Comparison: SID vs EID across the included studies.

**Figure 4:**
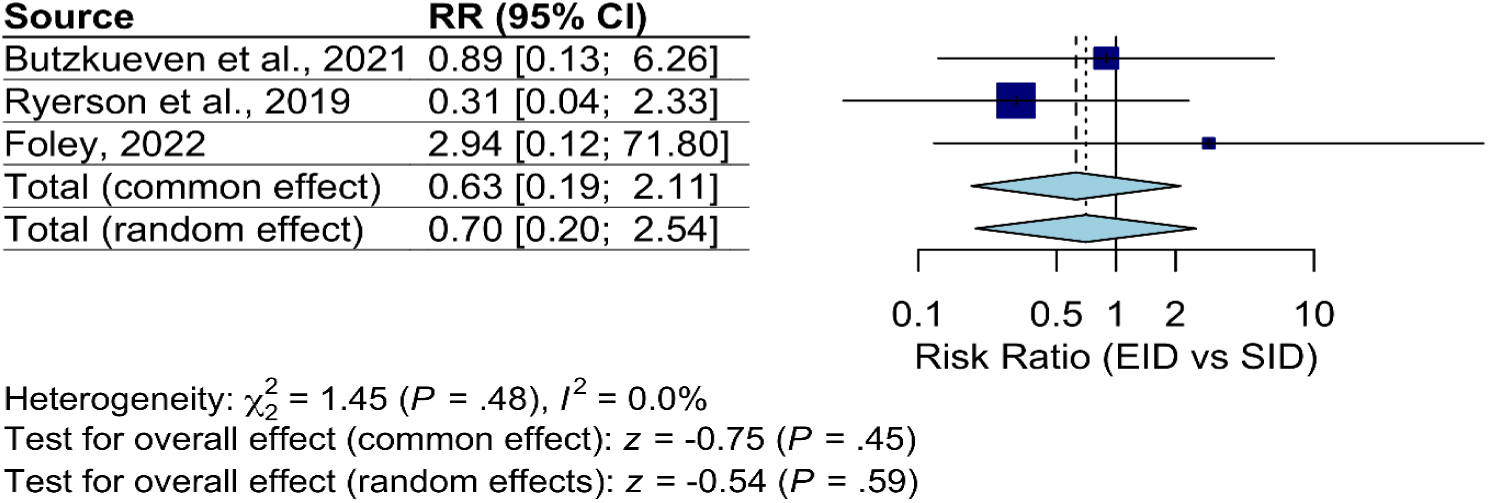
Forest Plot of PML Risk: SID vs EID (Main Analysis).

### 2.4 Search Strategy

Search strategies utilized a combination of controlled vocabulary (e.g., MeSH terms) and keywords related to “natalizumab,” “multiple sclerosis,” “extended interval dosing,” “standard interval dosing,” “cessation,” “relapse,” “disability progression,” and “PML.” Search filters limited results to human studies and English language, with a publication date from January 1, 2015. The search strategy was tailored for each database and is available on PROSPERO protocol registration.

### 2.5 Study Selection

Search results were imported into Zotero 6.0.30 for duplicate removal. Title/abstract screening and subsequent full-text assessment were performed independently by two reviewers (V.S. and M.B.) using Rayyan QCRI (17) . Conflicts were resolved through discussion or by a third reviewer (D.J.). The study selection process is detailed in the PRISMA flow diagram (15) (Figure 1).

### 2.6 Data Collection Process

Data extraction was conducted independently by two reviewers (V.S. and M.B.) using a standardized, piloted data extraction form in LibreOffice and Word. Disagreements were resolved by consensus or third-party adjudication (D.J.). Study authors were contacted for clarification or additional data when reported information was incomplete.

### 2.7 Data Items Extracted data included

- Study characteristics (design, country, funding, conflicts of interest).
- Participant characteristics (sample size, demographics, baseline disease characteristics).
- Intervention and comparator definitions (specific EID interval, cessation reason, post-cessation management).
- Outcome data for all pre-specified efficacy and safety endpoints, including effect estimates, measures of dispersion, and raw counts/denominators where available.

For cessation studies, where reported, data on post-cessation management strategies (e.g., immediate switch to another DMT vs. washout period) were extracted to assess heterogeneity. The following terms were used consistently and were not treated as interchangeable: relapse refers to a clinical relapse as defined by the primary study; disease reactivation refers to any clinical or MRI disease activity observed after cessation; and rebound refers to disease activity that a study explicitly defined as rebound, generally activity exceeding the pre-treatment baseline shortly after withdrawal. Study level definitions are provided in the results tables.

### 2.8 Risk of Bias in Individual Studies

The risk of bias was assessed independently by two reviewers (V.S. and M.B.).

- RCTs (n = 3) were evaluated using the Cochrane RoB 2.0 tool (19).
- Non-randomized studies (n = 24) were assessed using the ROBINS-I tool (20).
- Case series (n = 1) were appraised using the Joanna Briggs Institute (JBI) critical appraisal checklist (21).

Results are summarized graphically in Supplementary Figure 1A (RCTs) and Supplementary Figure 1B (non-randomized studies), with accompanying summary plots, generated using the robvis visualization tool (22). Detailed domain-level judgments for all studies are provided in Supplementary Table 1.

### 2.9 Synthesis Methods

Given that clinical decisions about dosing intervals cannot be separated from withdrawal planning in practice, we adopted a comprehensive approach examining both aspects of natalizumab management. This integrated perspective reflects real-world clinical decision-making where neurologists must consider the entire treatment continuum.

- Synthesis approach depended on clinical and methodological homogeneity. Meta-analysis used random-effects models for consistent outcomes. For rare events (PML), only studies reporting events were pooled. Person-year data were analyzed separately. Heterogeneity was quantified using I².
- Narrative synthesis followed SWiM guidelines (23) for outcomes with extreme heterogeneity. For ARR, meta-analysis was precluded due to EID definition variability (5 to 8 weeks), inconsistent reporting (7 calculation methods), and missing data. Median ARR difference was 0.00 (range −0.050 to +0.014).

For the post-cessation relapse analysis, we used a random-effects model and assessed heterogeneity using the I² statistic. To investigate sources of heterogeneity, we conducted pre-specified subgroup analyses based on cessation reason (planned vs. unplanned) and post-cessation management strategy (immediate switch vs. washout period). However, limited data reporting prevented formal subgroup analyses. The primary analysis included all studies, with sensitivity analysis excluding the outlier study (Weinstock-Guttman 2016) to assess robustness of findings. Supplementary Figure 3 shows the forest plot using Freeman-Tukey double arcsine transformation, and Supplementary Figure 4 presents the sensitivity analysis excluding the outlier study.

It is important to note that post-cessation management varied across studies; some cohorts included patients transitioning to alternative DMTs, while others involved washout periods. Outcomes were extracted irrespective of post-cessation treatment strategy, and this heterogeneity was considered in the interpretation of pooled relapse risk.

Sensitivity analyses assessed robustness. Evidence certainty was evaluated using GRADE (24) (Supplementary Table 4) and visualized using Critiplot (55) (Supplementary Figure 5).

### 2.10 Software

We used Zotero, Rayyan QCRI (17) , R statistical software (25) with the meta package (26), and LibreOffice/Word for data management and analysis.

## 3. Results

### 3.1 Study Selection

Our systematic search identified 1,338 records (1,316 from databases and 22 from trial registries), with an additional 12 records found through citation searching. After removing 297 duplicates, 1,041 titles and abstracts were screened, of which 818 were excluded for irrelevance. The 12 reports identified through citation searching entered the process at the retrieval stage and were not screened as records, which is why 1,041 rather than 1,053 records underwent title and abstract screening.

Reports sought for retrieval numbered 223 from databases and registries and 12 from citation searching. Nine reports could not be retrieved (7 from databases and registries, 2 from citation searching), leaving 216 reports from databases and registries and 10 reports from citation searching assessed for eligibility. Of these 226 reports, 198 were excluded (189 from databases and registries and 9 from citation searching) due to ineligible study design, population, intervention, or outcomes.

Ultimately, 28 studies met inclusion criteria (27 from databases and registries, 1 from citation searching). The PRISMA 2020 flow diagram (15) (Figure 1) summarizes the selection process, and excluded studies are listed in Supplementary Table 2.

### 3.2 Study Characteristics

The 28 included studies (2015–2025) comprised 3 randomized controlled trials (RCTs), 24 observational studies (prospective and retrospective cohorts), and 1 case series. Across all studies, the total sample size was 47,987 participants, although one large registry (Ryerson et al. (27), n=35,521) heavily influenced this number. Studies were conducted in single-country or multinational settings across Europe, North America, the Middle East, and Australia.

Two primary comparisons were addressed:

- Standard-Interval Dosing (SID) vs Extended-Interval Dosing (EID) of natalizumab, with EID definitions ranging from >5 weeks to 8-week intervals.
- Outcomes after cessation of natalizumab.

The key reported outcomes were annualized relapse rate (ARR), disability progression (EDSS worsening), NEDA-3, PML incidence, and post-cessation outcomes (relapse rates, time to relapse, and EDSS worsening). Funding and conflict of interest information were reported for 24 studies; 7 studies reported industry funding, largely from Biogen. Table 1 summarizes the detailed study characteristics, including the baseline demographics.

### 3.3 Risk of Bias

RCTs (n=3) were evaluated using ROB 2 (19): two were judged low risk, while one raised some concerns due to deviations from intended interventions.

Non-randomized studies (n=24) were assessed with ROBINS-I (20): 20 had moderate risk and 4 serious risk.

Confounding and missing data were the most common sources of bias. A case series (37) assessed with the JBI checklist was deemed moderate risk. Full risk-of-bias judgments are provided in Supplementary Table 1 and Supplementary Figures 1A and 1B.

### 3.4 Synthesis of Results

Feasibility of quantitative meta-analysis was pre-assessed for each outcome based on clinical and methodological homogeneity (Supplementary Table 3).

#### 3.4.1 SID vs EID: Efficacy Outcomes (ARR)

Eleven studies reported ARR related data, of which 9 provided data suitable for descriptive synthesis. ARR values were consistently low across both dosing strategies (unadjusted range: 0.0006 to 0.14). Based on the direction of point estimates, four studies reported a lower ARR with EID, three reported a lower ARR with SID, and two reported identical or nearly identical ARR between groups.

- The median ARR difference (EID – SID) was 0.000 (range –0.050 to +0.014).
- Pelle 2023 (45) was the only study with relapse counts and follow-up that allowed a formal comparison (risk ratio 0.44; 95% CI 0.16 to 1.26, based on patients with at least one relapse), showing no significant difference. Because EID intervals ranged from five to eight weeks across studies, we examined whether interval length was associated with the size or direction of the ARR difference; no significant association was found (Spearman rho = −0.36, p = 0.34), although the small number of studies limits this analysis.

Table 2A summarizes ARR outcomes. Meta-analysis was not possible due to heterogeneity in EID definitions, outcome reporting, and lack of raw event data.

Summary: Both SID and EID maintained very low relapse activity. No strategy consistently outperformed the other.

#### 3.4.2 SID vs EID: Disability Progression and NEDA-3

Heterogeneous outcome definitions and insufficient reporting prevented quantitative synthesis.

- Foley 2022 (34): 24 weeks of reported disability worsening: 92% [SID] vs 90% [EID] progression-free;
- NEDA-3: 163/242 [SID] vs 173/247 [EID].
- Pelle 2023 (45): Unconfirmed EDSS worsening: 13% [SID] versus 11% [EID].
- Riancho 2021 (46): Partial NEDA-3 reporting for EID only.

Table 2B presents study-specific outcomes for the SID versus EID comparison and for NEDA-3 during treatment; disability worsening following treatment discontinuation is reported separately in Table 2D.

Summary: Disability progression and NEDA-3 failure rates were modest across all regimens. Findings were broadly similar between groups, but the inconsistent outcome definitions and the small number of studies preclude any conclusion of equivalence.

#### 3.4.3 SID vs EID: Safety Outcome (PML)

Eight studies reported PML outcomes, one of them as person-years; Toorop et al. (50) did not report PML events and is therefore not listed in Table 2C. The meta-analysis of risk ratio (RR) included the three studies reporting PML events; the four studies reporting no events are presented in Table 2C for context (not included in pooled RR calculation).

- Prolonged exposure: in a TOUCH registry analysis of 35,521 anti-JC virus antibody positive patients, Ryerson et al. 2019 (27) demonstrated a 94% PML risk reduction with EID (adjusted HR 0.06, 95% CI 0.01 to 0.22). The person-year analysis of Ryerson et al. 2016 (48) (IRR 0.11; 95% CI 0.01 to 1.99) pointed in the same direction.
- Shorter exposure: Butzkueven et al. 2021 (30) (236 matched pairs, mean post-switch follow-up about 2 years) reported two PML cases in each group (210 unique SID patients, 236 EID patients). Foley et al. 2022 (34) (72 weeks) reported one asymptomatic PML case in EID (n=247) and none in SID (n=242).

Primary pooled analysis (3 studies with events): RR 0.70 (95% CI 0.20–2.54), I²=0%. Sensitivity analysis (excluding Foley 2022 asymptomatic case): RR 0.54 (95% CI 0.13–2.17). The pooled estimate was attenuated by inclusion of short-term studies with limited events and variable follow-up. Table 2C summarizes study-level PML data with exposure duration.

Summary: While the pooled analysis showed no significant difference in PML risk (RR 0.70), long-term data from Ryerson et al. (27) demonstrated substantial PML risk reduction with EID (94%) in patients with prolonged exposure. This discrepancy highlights limitations in meta-analyzing rare events without accounting for treatment duration. Ongoing monitoring remains warranted, particularly for long-term users.

Supplementary Figure 2: Forest Plot of PML Risk is also provided which includes Sensitivity Analysis Excluding Foley et al. 2022 (34).

#### 3.4.4 Outcomes After Cessation: Relapse and Disability

Four studies (n=861) reported the proportion of patients relapsing within 12 months post-cessation. A meta-analysis of these studies yielded a pooled relapse proportion of 41.7% (95% CI 30.6 to 53.6%) using a random-effects model (inverse variance with logit transformation). This estimate was largely driven by the Papeix et al. cohort (44) (n=715). Heterogeneity was observed (I²=66.9%, p=0.029), partially attributed to Weinstock-Guttman 2016 (59.3%). Sensitivity analysis excluding this study yielded a pooled estimate of 36.8% (95% CI 32.1 to 41.7%). A prediction interval of 21.8% to 64.7% indicates that the true proportion in a comparable future setting may plausibly lie anywhere within this range. Because reasons for cessation and post-cessation management differed across cohorts, this pooled proportion represents the observed risk within these study populations rather than a uniform individual risk for every patient discontinuing natalizumab in current practice.

Post-cessation management strategies varied across included studies. Some patients initiated another DMT immediately, some underwent a washout period with or without intravenous methylprednisolone, some restarted natalizumab (31% of the Papeix cohort), and some remained untreated (50% of the Papeix cohort). Reasons for cessation also differed (PML risk, planned interruption, or study protocol). Outcomes were extracted irrespective of post-cessation treatment; consequently, the pooled estimate reflects the average observed risk across these mixed pathways rather than the effect of any single strategy.

EDSS worsening rates after cessation were highly variable (19% to 100%) and were reported with different definitions, follow-up windows, and denominators; the two cohorts reporting EDSS-based rates gave 19% and 23%, one cohort reported secondary progression (18.0 to 22.7% by group) rather than EDSS worsening, and the 4-patient rebound case series (37) reported worsening in all patients. Because of this heterogeneity, no pooled or weighted estimate was calculated. Reported times to first relapse or rebound ranged from 3.0 to 6.0 months across the four studies reporting this outcome, clustering within the first 6 months; one cohort (32) separately reported time to MRI reactivation of 6.1 to 9.8 months.

A sensitivity analysis excluding Weinstock-Guttman 2016 showed minimal change in the pooled estimate (36.8%, 95% CI 32.1 to 41.7%), and leave-one-out analyses did not materially change the pooled proportion (Supplementary Table 5), suggesting robustness of findings despite the heterogeneity introduced by this study’s design.

Forest plots for the primary analysis, Freeman-Tukey transformation, and sensitivity analysis are presented in Figure 5, Supplementary Figure 3, and Supplementary Figure 4, respectively.

**Figure 5.**
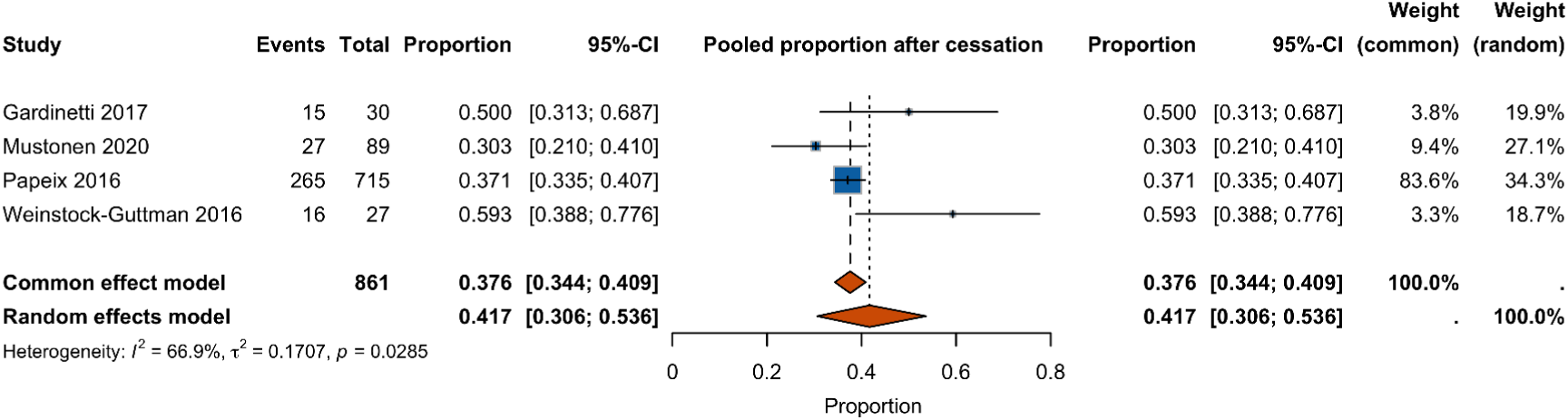
Forest plot of the proportion of patients relapsing within 12 months after natalizumab cessation (random-effects model).

**Figure 6.**
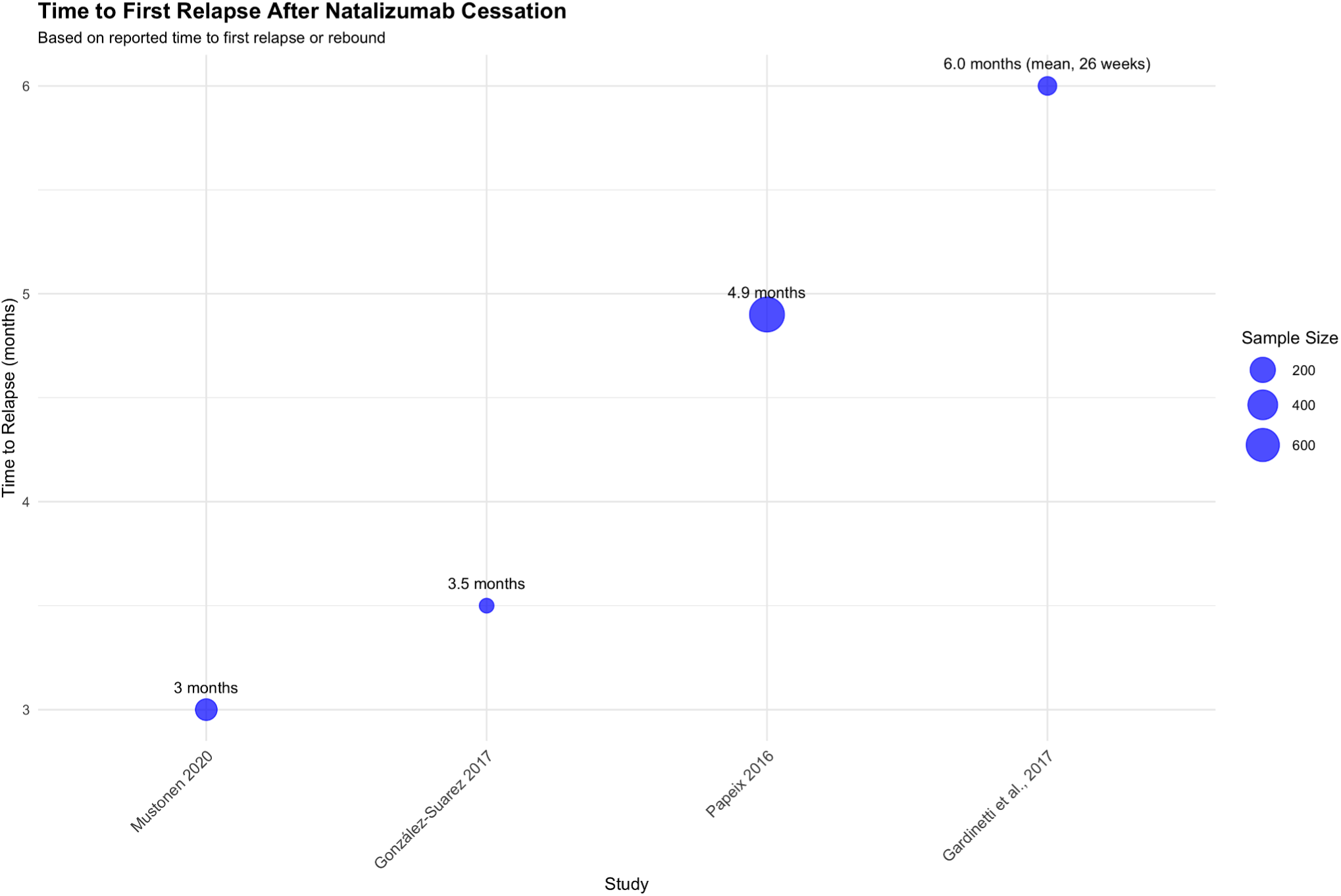
Time to first relapse or rebound across the four studies reporting this outcome (Papeix: time by which 25% of patients had relapsed; Gardinetti: mean; Mustonen: median within the rebound subgroup; González-Suarez: mean time to rebound). One further cohort reported time to MRI reactivation separately (Table 2D).

**Figure 7.**
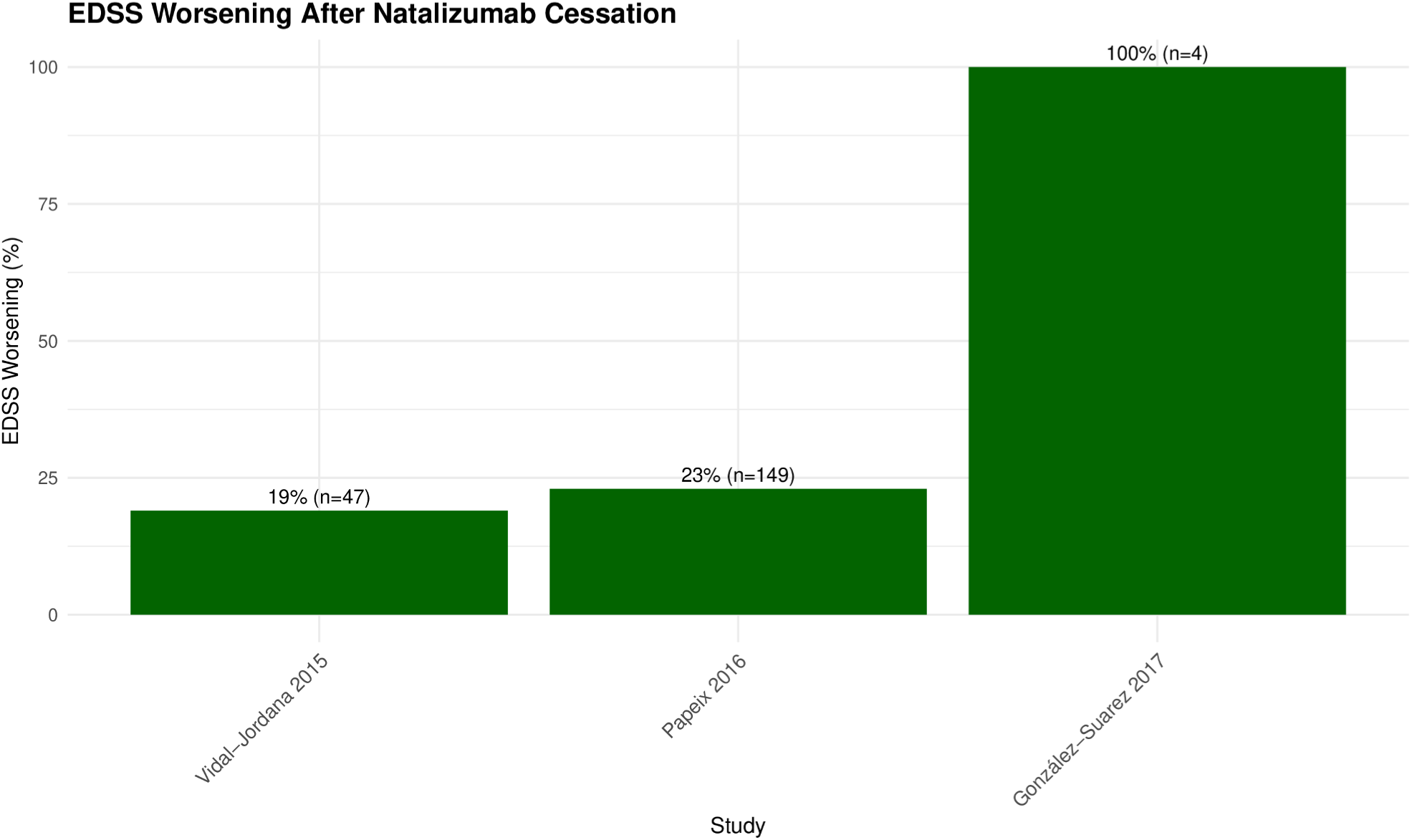
Bar plot of EDSS worsening rates after cessation in the three studies reporting this outcome with EDSS-based measures. No pooled estimate was calculated; the 4-patient rebound case series (37) reported worsening in all patients. Coerver et al. reported secondary progression (18.0 to 22.7% by group) rather than EDSS worsening and is therefore not shown.

Table 2D summarizes cessation outcomes.

Summary: Natalizumab cessation is associated with a relapse risk of 41.7% at 12 months based on the pooled analysis of 4 studies, with most relapses occurring early. This finding indicates a high rate of disease reactivity following treatment discontinuation.

Forest plot of pooled relapse rate after natalizumab cessation using inverse variance method with logit transformation (random-effects model). Squares represent individual study proportions with size proportional to sample weight. Horizontal lines represent 95% confidence intervals. Diamond represents pooled estimate (41.7%, 95% CI 30.6 to 53.6%).

Supplementary Figure 3. Forest plot using Freeman-Tukey double arcsine transformation for relapse risk after natalizumab cessation. Squares represent individual study proportions with size proportional to sample weight. Horizontal lines represent 95% confidence intervals. Diamond represents pooled estimate (41.5%, 95% CI 30.1 to 53.4%).

Supplementary Figure 4. Sensitivity analysis forest plot excluding Weinstock-Guttman 2016. Squares represent individual study proportions with size proportional to sample weight. Horizontal lines represent 95% confidence intervals. Diamond represents pooled estimate (36.8%, 95% CI 32.1 to 41.7%).

### 3.5 Sensitivity Analyses

Leave-one-out analyses confirmed robustness of pooled results for both PML risk and cessation relapse rates. Exclusion of single studies influenced heterogeneity but not overall interpretation (Supplementary Table 5).

### 3.6 Reporting Bias

Publication bias could not be formally assessed as each outcome had fewer than 10 studies, insufficient for funnel plots or statistical testing.

### 3.7 Certainty of Evidence (GRADE)

Certainty of evidence for each outcome is summarized in Supplementary Table 4 and visualised in Supplementary Figure 5:

- SID vs EID:
- `o` ARR = Low (risk of bias, inconsistency)
- `o` Disability progression = Very Low (bias, inconsistency, indirectness)
- `o` PML = Low (bias, imprecision)
- `o` NEDA-3 = Low (bias, imprecision, single study)
- Outcomes After Cessation:
- `o` Relapse = Low (bias, heterogeneity)

### Time to first relapse = Low (risk of bias, inconsistency)

- EDSS worsening = Very Low (bias, inconsistency, imprecision)

*The registry analysis of prolonged exposure (94% reduction in Ryerson 2019) provides the strongest available evidence for PML risk reduction; the pooled estimate across mixed exposure durations remains imprecise (see Supplementary Table 4)*.

## 4. Discussion

### 4.1. Interpreting the Results

This systematic review provides a systematic analysis of natalizumab treatment outcomes. Our analysis of post-cessation outcomes, based on 4 studies (n=861), revealed that 41.7% (95% CI 30.6 to 53.6%) of patients in the included cohorts experience relapse within 12 months of discontinuing natalizumab; because post-cessation management and reasons for cessation differed across cohorts, this figure describes the observed risk in these populations rather than a uniform individual risk. This estimate was robust in sensitivity analysis excluding an outlier study (36.8%; 95% CI 32.1 to 41.7%). The heterogeneity observed (I²=66.9%) was largely driven by one study (Weinstock-Guttman 2016) and differences in post-cessation management strategies across cohorts. The observed relapse risk highlights the challenges associated with treatment discontinuation.

Regarding dosing strategies, our analysis found no consistent difference in efficacy between SID and EID. Across nine studies, the median ARR difference was zero (range −0.050 to +0.014); these descriptive findings do not establish equivalence or non-inferiority, which would require pre-specified margins and formal testing. Similarly, when disability progression and NEDA-3 results were reported, they were modest and similar between groups, despite the limitations of different classifications.

The safety outcomes, specifically PML incidence, were meta-analyzed across the three studies that recorded occurrences. The pooled risk ratio was 0.70 (95% confidence interval: 0.20–2.54). While our point estimate shows that EID may reduce PML risk, the confidence intervals are broad and can account for both clinically significant benefit and harm. The frequency of PML episodes, combined with the observational nature of the evidence, limits the certainty of this finding. Our pooled PML estimate (RR 0.70) differs from Ryerson 2019’s 94% reduction because meta-analysis of rare events is highly sensitive to study duration. In patients with prolonged exposure, the registry analysis showed substantial PML risk reduction with EID, supporting its use in JCV-positive patients with long treatment duration.

### 4.2. Treatment Continuum Framework

Our findings support examining natalizumab as a treatment continuum because: (1) dosing strategy selection affects future cessation planning, (2) treatment duration impacts both PML risk and post-cessation relapse risk, and (3) neurologists must consider both aspects simultaneously for optimal patient management. This approach aligns with clinical practice where treatment decisions cannot be made in isolation.

### 4.3. Limitations of the Evidence

Key limitations include: (1) Predominance of observational studies (24/28) with potential for confounding despite adjustment attempts, and inclusion of three cohorts with mixed populations (RRMS together with progressive MS) in full as published; (2) Substantial heterogeneity in EID definitions (5 to 8 weeks), where intervals of five and eight weeks do not represent the same exposure, and heterogeneous outcome reporting precluding meta-analysis for efficacy outcomes; (3) Imprecise PML estimates due to rare events, with one large registry (Ryerson 2019, n=35,521) dominating findings; (4) Limited data on optimal post-cessation management strategies, with most studies reporting observational outcomes rather than comparative effectiveness of different approaches; (5) Potential residual confounding despite statistical adjustments in observational studies

Regarding our comprehensive scope examining both dosing and withdrawal outcomes, we recognize that some may question whether these topics belong in a single review. However, we believe this integrated approach is justified because: (1) clinical decisions about dosing intervals cannot be made without considering future withdrawal implications, (2) withdrawal outcomes depend on prior dosing history, and (3) neurologists managing natalizumab treatment must consider the entire treatment continuum. This comprehensive perspective reflects real-world clinical practice and provides the integrated evidence needed for clinical decision-making.

### 4.4. Limitations of the Review Procedure

Our review followed PRISMA 2020 guidelines (15). However, several limitations should be acknowledged. The search was restricted to studies published after 2015 and in English, which may have induced bias. Due to heterogeneity in outcome definitions, narrative synthesis was required for the majority of efficacy outcomes. Finally, the small number of papers for each outcome limited our formal assessment of publication bias. The inclusion of one study from late 2014 (published in 2015) is a minor change from the protocol, stated for transparency.

### 4.5 Implications for Practice and Research

Clinical implications: Based on our findings, we propose specific clinical considerations for natalizumab management: • For treatment initiation: Consider the entire treatment continuum from the outset, including eventual discontinuation planning. • For ongoing treatment: For stable patients considering EID, discuss that while efficacy may be maintained, managing discontinuation remains a challenge regardless of dosing strategy. • For discontinuation planning: The 41.7% relapse risk within 12 months in the included cohorts, with most events occurring within the first 6 months, suggests that clinicians should carefully plan the timing of alternative therapies. While this review did not compare specific transition protocols, the high relapse rate supports minimizing the time without effective treatment. • For patient counseling: Inform patients that a significant proportion experience relapse after stopping natalizumab, with most events occurring in the first 6 months. • For high-risk patients: Those with higher relapse risk prior to natalizumab or longer treatment duration may require particularly close monitoring.

Future research should prioritize: (1) Randomized trials comparing different post-cessation management strategies, including optimal timing for initiating alternative therapies; (2) Prospective studies identifying risk factors for post-cessation relapse to develop prediction models; (3) Studies evaluating cost-effectiveness of different monitoring and transition strategies; (4) Research on patient-reported outcomes and preferences regarding natalizumab discontinuation planning; (5) Long-term follow-up studies assessing disability outcomes beyond 12 months post-cessation. Additionally, large prospective registries are needed to better define risk factors for post-cessation relapse. While further research on EID safety is warranted, a significant knowledge gap remains in understanding how to safely manage the period following natalizumab discontinuation.

## 5. Conclusion

This review identifies a high relapse risk following natalizumab cessation (41.7% at 12 months in the included cohorts), with sensitivity analysis confirming a 36.8% risk after excluding an outlier study. No consistent efficacy difference between EID and SID was identified, although formal equivalence was not tested; neurologists should adopt a treatment continuum approach that integrates dosing decisions with cessation planning. Future research should prioritize optimal transition protocols to mitigate this high-risk period.

Clinical Practice Summary: The pooled relapse risk of 41.7% (95% CI 30.6 to 53.6%) within 12 months of natalizumab cessation in the included cohorts necessitates careful treatment planning; this figure reflects mixed post-cessation pathways and should not be taken as the individual risk for every patient. Sensitivity analysis showed a 36.8% relapse risk (95% CI 32.1 to 41.7%) when excluding an outlier study. The first 6 months post-cessation represent a high-risk period, with reported times to relapse or rebound ranging from 3.0 to 6.0 months. Regarding dosing strategies, the available evidence did not identify a consistent efficacy difference between EID and SID, and formal equivalence was not tested; treatment decisions should consider the entire continuum from initiation through discontinuation. While this review did not analyze specific transition therapies, the observed relapse rate supports the general principle of minimizing treatment gaps.

## Data Availability

All data analyzed in this study are extracted from previously published studies and are included in the manuscript and Supplementary Files.

## Acknowledgments

Not applicable.

## Ethics Approval and Consent to Participate

Not applicable. This study is a systematic review and meta-analysis of previously published data.

## Consent for Publication

Not applicable.

## Availability of Data and Materials

All data analyzed in this study were obtained from previously published sources. No new datasets were generated or analyzed during the current study.

## Author Contributions

V.S. conceptualized and designed the study, developed the protocol (PROSPERO registration), conducted the literature search with assistance from M.B., performed title/abstract screening and full-text assessment as first reviewer with M.B. as second reviewer, extracted data, conducted formal analyses including meta-analyses and GRADE assessment, interpreted the results, drafted the entire manuscript including all sections, figures, and tables, created all visualizations (forest plots and PRISMA flowchart), and provided project administration, supervision, and correspondence as the corresponding author. M.B. assisted with literature search, served as second reviewer for title/abstract screening and full-text assessment, performed risk of bias assessment for all included studies using Cochrane RoB 2.0 for RCTs and ROBINS-I for observational studies, served as third-party adjudicator for conflicts during screening and data extraction, and provided critical review of the final manuscript. D.J. assisted with data extraction as secondary reviewer, served as third-party conflict resolver during study selection, and provided critical review of the final manuscript. All authors approved the final version of the manuscript and agree to be accountable for the work’s integrity and accuracy.

