## Supplementary Files for "Natalizumab Treatment Continuum in RRMS: A Systematic Review and Meta-Analysis of Cessation Risks and Dosing Strategies"

**Supplementary Table 1: Risk of Bias Assessment**

**Risk of Bias Assessment of Included RCTs (ROB 2 Tool)**

| **Author, Year** | **Randomization** | **Deviations** | **Missing Data** | **Measurement** | **Reporting** | **Overall RoB** |
|---|---|---|---|---|---|---|
| Kaufman (RESTORE), 2015 | Low | Some concerns | Low | Low | Low | Low |
| Foley et al., 2022 (NOVA) | Low | Low | Low | Low | Low | Low |
| Weinstock-Guttman et al., 2016 | Some concerns | High | Low | Some concerns | Low | Some concerns |

**Risk of Bias Assessment of Non-Randomized Studies (ROBINS-I Tool)**

| **Author, Year** | **Confounding** | **Selection** | **Classification** | **Deviations** | **Missing data** | **Measurement** | **Reporting** | **Overall RoB** |
|---|---|---|---|---|---|---|---|---|
| Ryerson, 2016 | Moderate | Low | Low | Low | Low | Moderate | Low | Moderate |
| Ruggieri, 2019 | Moderate | Low | Low | Low | Moderate | Low | Low | Moderate |
| Butzkueven, 2020 | Moderate | Moderate | Low | Moderate | Moderate | Low | Low | Moderate |
| Toorop, 2024 | Serious | Low | Low | Moderate | Moderate | Low | Low | Serious |
| Al-Mahdawi, 2023 | Moderate | Low | Low | Low | Moderate | Low | Low | Moderate |
| Coerver, 2021 | Moderate | Low | Low | Low | Moderate | Low | Low | Moderate |
| Clerico, 2020 | Moderate | Low | Low | Low | Moderate | Low | Low | Moderate |
| Gardinetti, 2017 | Serious | Moderate | Low | Moderate | Low | Moderate | Moderate | Serious |
| Jeantin, 2023 | Moderate | Low | Low | Low | Moderate | Low | Moderate | Moderate |
| Riancho, 2021 | Serious | Moderate | Low | Serious | Moderate | Low | Moderate | Serious |

| | | | | | | | | |
|---|---|---|---|---|---|---|---|---|
| Yamout, 2018 | Serious | Moderate | Low | Low | Low | Moderate | Low | Moderate |
| Ruggieri, 2024 | Moderate | Serious | Low | Moderate | Moderate | Low | Low | Moderate |
| Bernardes et al., 2024 | Moderate | Low | Low | Low | Low | Moderate | Low | Moderate |
| De Mercanti et al., 2021 | Moderate | Low | Low | Low | Moderate | Serious | Low | Moderate |
| Fuentes-Rumí et al., 2020 | Moderate | Low | Low | Low | Moderate | Moderate | Low | Moderate |
| Haas et al., 2017 | Moderate | Moderate | Low | Low | Low | Low | Low | Moderate |
| Jakimovski et al., 2023 | Moderate | Low | Low | Low | Moderate | Low | Low | Moderate |
| Kister et al., 2018 | Moderate | Moderate | Low | Low | Moderate | Low | Low | Moderate |
| Mustonen et al., 2020 | Moderate | Low | Low | Low | Moderate | Low | Low | Moderate |
| Papeix et al., 2016 | Moderate | Moderate | Low | Low | Serious | Low | Low | Moderate |
| Pelle et al., 2023 | Moderate | Low | Low | Moderate | Serious | Low | Low | Serious |
| van Kempen et al., 2020 | Moderate | Low | Low | Low | Low | Low | Low | Moderate |
| Vidal-Jordana et al., 2015 | Moderate | Low | Low | Moderate | Moderate | Low | Low | Moderate |
| Ryerson et al 2019 | Moderate | Moderate | Low | Low | Low | Low | Low | Moderate |

**JBI (Case Series)**

| Author | Clear eligibility criteria? | Condition measured consistently? | Valid methods used for identification? | Consecutive recruitment used? | Complete outcomes reported? | Follow-up long enough? | Statistical methods appropriate? | Demographics detailed? | Clinical info and interventions described | Adverse events reported? | Overall risk |
|---|---|---|---|---|---|---|---|---|---|---|---|
| González-Suarez et al., 2017 | Yes | Yes | Yes | No | Yes | Yes | Yes | Yes | Yes | Yes | Moderate |

Note: Overall risk of bias judgments represent the integrated assessment of the two reviewers; a serious rating in a single domain did not automatically determine the overall judgment when that domain was judged less critical for the reported outcomes.

**Supplementary Table 2: Excluded papers.**

**Total count overall including exclusion from the Database/registers and citation records**

| **Reason for Exclusion (overall)** | **Number of Studies (n)** |
|---|---|
| Findings without clinical endpoints | 29 |
| Studies that did not compare SID vs EID or cessation vs continuation | 71 |
| Wrong study design | 51 |
| Other DMTs as intervention | 36 |
| Wrong population | 9 |
| Wrong intervention | 2 |

To check exact overall exclusion numbers seperately for Databases/Registers and Citation Records, please check the PRISMA flowchart in the Manuscript.

| Authors | Title | Year | Reason for Exclusion | Clarification |
|---|---|---|---|---|
| Brandstadter et al. | The use of natalizumab for multiple sclerosis | 2017 | Studies not comparing SID vs EID or cessation vs continuation | Narrative review article on natalizumab's efficacy and safety; does not contain a comparative study of dosing intervals or cessation. |
| Doshi et al. | Regional Efficacy of Natalizumab Treatment in Relapsing-Remitting Multiple Sclerosis (RRMS) | 2015 | Findings without clinical endpoints | Conference abstract; lacks detailed reporting of ARR, EDSS, or comparator data needed for analysis. |
| Evdoshenko et al. | Real-world study of efficacy, risk management and reasons for discontinuation of natalizumab for treatment of multiple sclerosis in Russia. | 2019 | Studies not comparing SID vs EID or cessation vs continuation | Observational study on reasons for discontinuation; does not compare outcomes between a SID and an EID group or a continuation vs cessation group. |
| Guger et al. | Pregnancy Outcomes in Patients With Multiple Sclerosis Exposed to Natalizumab—A Retrospective Analysis From the Austrian Multiple Sclerosis Treatment Registry | 2020 | Wrong study design | Study focuses exclusively on pregnancy outcomes, not on the comparative effectiveness of different natalizumab dosing schedules or cessation. |
| Correia et al. | Predictors of first-line treatment persistence in a Portuguese cohort of relapsing-remitting multiple sclerosis | 2016 | Other DMTs as intervention | Study investigates predictors of persistence on first-line DMTs (interferon/glatiramer acetate), not natalizumab dosing or cessation. |
| Dalla Costa et al. | Neuroaxonal damage in natalizumab-treated MS patients: The role of JCV antibody titres | 2024 | Findings without clinical endpoints | Study investigates a biomarker (sNfL) in relation to JCV status; does not report comparative ARR or EDSS outcomes for |

| | | | | |
|---|---|---|---|---|
| | | | | dosing intervals/cessation. |
| Delbue et al. | Natalizumab treatment of multiple sclerosis: new insights | 2017 | Studies not comparing SID vs EID or cessation vs continuation | Narrative review article summarizing insights on natalizumab use and PML risk; not a primary study comparing dosing or cessation. |
| de Oliveira et al. | Natalizumab treatment in multiple sclerosis: the experience from two Brazilian MS centers | 2015 | Studies not comparing SID vs EID or cessation vs continuation | Single-arm cohort study describing outcomes on natalizumab; lacks a comparator group (EID or cessation). |
| Schwehr et al | Informing Medication Discontinuation Decisions among Older Adults with Relapsing-Onset Multiple Sclerosis | 2020 | Studies not comparing SID vs EID or cessation vs continuation | Completely off from the PICO criteria |
| Iaffaldano et al. | Natalizumab reduces serum pro-angiogenic activity in MS patients | 2018 | Findings without clinical endpoints | Mechanistic study measuring angiogenic activity and osteopontin levels; does not report clinical endpoints like ARR or EDSS. |
| Capra et al. | Natalizumab is associated with early improvement of working ability in relapsing-remitting multiple sclerosis patients: WANT observational study results | 2021 | Studies not comparing SID vs EID or cessation vs continuation | Single-arm study assessing work ability and quality of life outcomes on natalizumab; no comparator group for dosing or cessation. |
| Fox et al. | MS disease activity in RESTORE: a randomized 24-week natalizumab treatment interruption study | 2014 | Wrong study design | Study published in 2014, which is outside the inclusion window (2015 or later). |
| Guger et al. | Long-term outcome and predictors of long-term disease activity in natalizumab-treated patients with multiple sclerosis: real life data from the Austrian MS Treatment | 2021 | Studies not comparing SID vs EID or cessation vs continuation | Single-arm, long-term observational study on natalizumab; does not contain a comparison of different dosing intervals or cessation. |
| Foley et al. | Long-term natalizumab treatment is associated with sustained improvements in quality of life in patients with multiple sclerosis | 2017 | Studies not comparing SID vs EID or cessation vs continuation | Single-arm study assessing quality of life outcomes on natalizumab; no comparator group for dosing or cessation. |
| Boziki et al. | Long-Term Efficacy Outcomes of Natalizumab vs. Fingolimod in | 2021 | Other DMTs as intervention | Comparator is fingolimod, a different DMT. The study does |

| | | | | |
|---|---|---|---|---|
| | Patients With Highly Active Relapsing-Remitting Multiple Sclerosis: Real-World Data From a Multiple Sclerosis Reference Center | | | not compare natalizumab SID vs EID or continuation vs cessation. |
| Chisari et al. | Long-term effectiveness of natalizumab in secondary progressive multiple sclerosis: A propensity-matched study | 2024 | Wrong study design | Population is patients with Secondary Progressive MS (SPMS), not Relapsing-Remitting MS (RRMS). |
| Bigaut et al. | Long-term effect of natalizumab in patients with RRMS: TYSTEN cohort | 2021 | Studies not comparing SID vs EID or cessation vs continuation | Single-arm, long-term observational study on natalizumab; does not contain a comparison of different dosing intervals or cessation. |
| Butzkueven | Long-term clinical outcomes in patients with multiple sclerosis who are initiating disease-modifying therapy with natalizumab compared with BRACETD first-line therapies | 2024 | Other DMTs as intervention | Comparator is first-line therapies (BRACETD), not a natalizumab dosing interval or cessation strategy. |

| | | | | |
|---|---|---|---|---|
| Auer et al. | Experiences in treatment of multiple sclerosis with natalizumab from a real-life cohort over 15 years | 2021 | Studies not comparing SID vs EID or cessation vs continuation | Single-arm cohort study describing long-term outcomes and factors for rebound; does not compare SID vs EID or include a cessation vs continuation group. |
| Avila et al. | Evaluating natalizumab first-line and later-line use in multiple sclerosis: a US claims database analysis | 2025 | Other DMTs as intervention | Study compares natalizumab as 1st-line vs 2nd-line therapy; the comparator is not a dosing interval or cessation strategy. |
| Janoschka et al. | Enhanced pathogenicity of Th17 cells due to natalizumab treatment: Implications for MS disease rebound | 2023 | Findings without clinical endpoints | Mechanistic laboratory study investigating Th17 cell biology; does not report clinical outcomes like ARR or EDSS. |
| Granella et al. | Eligibility Criteria to Natalizumab Therapy in Patients with Relapsing-remitting Multiple Sclerosis: A Real-life Study in an Italian Population-based Cohort | 2015 | Studies not comparing SID vs EID or cessation vs continuation | Study describes baseline characteristics of patients prescribed natalizumab; not an analysis of outcomes based on dosing or cessation. |
| Cobo-Calvo et al. | Effectiveness of natalizumab in patients with highly active relapsing | 2015 | Studies not comparing SID | Single-arm study assessing effectiveness in a highly active |

| | | | | |
|---|---|---|---|---|
| | remitting multiple sclerosis | | vs EID or cessation vs continuation | subgroup; lacks a comparator group for dosing intervals or cessation. |
| Fagius et al. | Discontinuation of disease modifying treatments in middle aged multiple sclerosis patients. First line drugs vs natalizumab | 2017 | Other DMTs as intervention | Study compares discontinuation of first-line DMTs vs natalizumab; does not compare natalizumab SID vs EID or cessation vs continuation. |
| Jacques et al. | Cognitive evolution in natalizumab-treated multiple sclerosis patients | 2016 | Studies not comparing SID vs EID or cessation vs continuation | Single-arm study assessing cognitive outcomes on natalizumab; no comparator group for dosing intervals or cessation. |
| Carotenuto et al. | CD4/CD8 ratio during natalizumab treatment in multiple sclerosis patients | 2017 | Findings without clinical endpoints | Laboratory study monitoring lymphocyte ratios; does not report comparative clinical endpoints (ARR, EDSS) for dosing or cessation. |
| Cohen et al. | BEST-MS: A prospective head-to-head comparative study of natalizumab and fingolimod in active relapsing MS | 2021 | Other DMTs as intervention | Comparator is fingolimod, a different DMT. The study does not compare natalizumab SID vs EID or continuation vs cessation. |
| Kornek | An update on the use of natalizumab in the treatment of multiple sclerosis: appropriate patient selection and special considerations | 2015 | Studies not comparing SID vs EID or cessation vs continuation | Narrative review article discussing patient selection; not a primary study comparing dosing intervals or cessation outcomes. |
| Iaffaldano et al. | A comparison of natalizumab and ocrelizumab on disease progression in multiple sclerosis | 2024 | Other DMTs as intervention | Comparator is ocrelizumab, a different DMT. The study does not compare natalizumab SID vs EID or continuation vs cessation. |
| Andersen et al. | The effectiveness of natalizumab vs fingolimod–A comparison of international registry studies | 2021 | Other DMTs as intervention | Comparator is fingolimod, a different DMT. The study is a meta-analysis of registry studies comparing two different drugs, not natalizumab dosing strategies. |
| Correia et al. | Long-term effectiveness and safety of natalizumab in a Portuguese | 2017 | Studies not comparing SID | Single-arm, long-term observational study on |

| | | | | |
|---|---|---|---|---|
| | population | | vs EID or cessation vs continuation | natalizumab; does not contain a comparison of different dosing intervals or cessation. |
| Deslandes et al. | Effectiveness and Adverse Events of Use of Natalizumab in a Brazilian Cohort of Patients With Multiple Sclerosis | 2020 | Studies not comparing SID vs EID or cessation vs continuation | Single-arm cohort study describing effectiveness and safety in a Brazilian population; lacks a comparator group for dosing or cessation. |
| Hemmer et al. | Efficacy and Safety of Proposed Biosimilar Natalizumab (PB006) in Patients With Relapsing-Remitting Multiple Sclerosis: The Antelope Phase 3 Randomized Clinical Trial | 2023 | Wrong study design | Study compares biosimilar vs reference natalizumab, both administered on a standard 4-week schedule. It does not compare different dosing intervals (SID vs EID) or cessation. |
| Hartung et al. | TRUST - A Study to Evaluate an Integrated Approach for Optimized Patient Management in Multiple Sclerosis Patients Treated with Natalizumab - First Analysis of Baseline Data (P2.147) | 2016 | Wrong study design | Conference abstract describing the design and baseline data of an observational study; does not report outcomes related to dosing intervals or cessation. |

| | | | | |
|---|---|---|---|---|
| Giovannoni et al. | Sustained Disability Improvement as Assessed by a Multicomponent Endpoint in Secondary Progressive Multiple Sclerosis (SPMS) Patients: A Post Hoc Analysis from ASCEND (P5.359) | 2017 | Wrong study design | Population is patients with Secondary Progressive MS (SPMS), not Relapsing-Remitting MS (RRMS). The analysis is also from a trial where the primary endpoint was not met. |
| Chen et al. | Superior effects of natalizumab versus other DMTs on patient-reported outcomes in people with multiple sclerosis | 2022 | Other DMTs as intervention | Comparator is a pooled group of "other DMTs". The study does not compare natalizumab SID vs EID or continuation vs cessation. |
| Fuchs et al. | Natalizumab for the Treatment of Relapsing- | 2019 | Studies not comparing | Appears to be a general review or summary article on natalizumab; |

| | | | | |
|---|---|---|---|---|
| | Remitting Multiple Sclerosis | | SID vs EID or cessation vs continuation | not a primary study comparing dosing intervals or cessation outcomes. |
| Efthimios et al. | Long-Term Effectiveness of Natalizumab in Patients with Relapsing-Remitting Multiple Sclerosis Treated in the Routine Care in Greece: Results from the Multicenter, Observational 5-Year Prospective Study 'TOPICS Greece' | 2021 | Studies not comparing SID vs EID or cessation vs continuation | Single-arm, long-term observational study on natalizumab; does not contain a comparison of different dosing intervals or cessation. |
| Gudesblatt et al. | Improvement in Cognitive Function as Measured by NeuroTrax in Patients with Relapsing Multiple Sclerosis Treated with Natalizumab: A 2-Year Retrospective Analysis | 2018 | Studies not comparing SID vs EID or cessation vs continuation | Single-arm study assessing cognitive outcomes on natalizumab; no comparator group for dosing intervals or cessation. |
| Hersh et al. | Impact of natalizumab on quality of life in a real-world cohort of patients with multiple sclerosis: Results from MS PATHS | 2021 | Studies not comparing SID vs EID or cessation vs continuation | Single-arm study assessing quality of life outcomes on natalizumab; no comparator group for dosing intervals or cessation. |
| Diouf et al. | Effectiveness of multiple disease-modifying therapies in relapsing-remitting multiple sclerosis: causal inference to emulate a multiarm randomised trial | 2023 | Other DMTs as intervention | Study compares natalizumab against multiple other DMTs (fingolimod, dimethyl fumarate, etc.). The comparator is not a natalizumab dosing interval or cessation strategy. |
| Hartung et al. | ASCEND Phase 3 Trial Open-Label Extension Study Results: Natalizumab May Delay Disability Progression in Secondary Progressive Multiple Sclerosis (SPMS) (P5.330) | 2017 | Wrong study design | Population is patients with Secondary Progressive MS (SPMS), not Relapsing-Remitting MS (RRMS). |
| Guerra et al. | Long-term comparative analysis of no evidence of disease activity (NEDA-3) status between multiple sclerosis patients treated with | 2021 | Other DMTs as intervention | Comparator is fingolimod, a different DMT. The study does not compare natalizumab SID vs EID or continuation vs cessation. |

| | | | | |
|---|---|---|---|---|
| | natalizumab and fingolimod for up to 4 years | | | |
| DeLuca | Exploring multiple sclerosis: Cognitive challenges, therapeutic advances, and comprehensive care insights | 2022 | Wrong study design | Appears to be a news article or commentary summarizing the NOVA trial results, not a primary study. |
| Guger et al. | De□Escalation Treatment Strategies From Natalizumab in Patients With Relapsing Multiple Sclerosis in Austria | 2025 | Studies not comparing SID vs EID or cessation vs continuation | Study analyzes outcomes after switching from natalizumab to other DMTs; it does not compare natalizumab SID vs EID or include a continuation group. |
| Jamroz-Wiśniewska et al. | Modified Rio Score with Platform Therapy Predicts Treatment Success with Fingolimod and Natalizumab in Relapsing-Remitting Multiple Sclerosis Patients | 2021 | Other DMTs as intervention | Study focuses on predicting response to fingolimod and natalizumab. It does not compare natalizumab dosing strategies; the groups are based on the drug chosen, not its dosing schedule. |
| Beume et al. | Massive exacerbation of multiple sclerosis after withdrawal and early restart of treatment with natalizumab | 2015 | Studies not comparing SID vs EID or cessation vs continuation | Case report describing a rebound event; does not provide a comparative analysis of outcomes between a cessation group and a continuation group. |
| Bringeland et al. | Wearing-off at the end of natalizumab dosing interval and risk of MS disease activity: A prospective 1-year follow-up study | 2020 | Studies not comparing SID vs EID or cessation vs continuation | Study investigates "wearing-off" symptoms within the standard 4-week cycle; it does not compare outcomes between a SID group and an EID group. |
| Cobo-Calvo et al. | Leukocyte adhesion molecule dynamics after Natalizumab withdrawal in Multiple Sclerosis | 2016 | Findings without clinical endpoints | Laboratory study measuring biomarker dynamics after withdrawal; does not report comparative clinical outcomes (ARR, EDSS) for a cessation vs continuation group. |

| | | | | |
|---|---|---|---|---|
| Cordioli et al. | Early detection and favourable outcome of natalizumab-related progressive multifocal leukoencephalopathy (PML) in two multiple sclerosis patients | 2015 | Wrong study design | Case series focused on PML diagnosis and management; does not investigate natalizumab dosing intervals or comparative cessation outcomes. |
| Derfuss et al. | α4-integrin receptor desaturation and disease activity return after | 2017 | Findings without clinical | Pharmacodynamic study measuring receptor saturation |

| | | | | |
|---|---|---|---|---|
| | natalizumab cessation | | endpoints | as a biomarker; clinical relapse data is secondary and not compared against a continuation group. |
| Dhanapalaratnam et al. | 5 Neuroprotective effects of metformin in diabetic peripheral neuropathy | 2024 | Wrong study design | Study is on diabetic peripheral neuropathy and an unrelated drug (metformin); completely outside the scope of this review. |
| Ferrè et al. | Recurrence of disease activity after repeated Natalizumab withdrawals | 2015 | Studies not comparing SID vs EID or cessation vs continuation | Case series describing rebound after withdrawal; does not provide a comparative analysis against a continuation group. |
| Fiander et al. | Clinical Course of Relapsing Remitting Multiple Sclerosis Post-Natalizumab | 2019 | Studies not comparing SID vs EID or cessation vs continuation | Single-arm study describing outcomes after switching from natalizumab to fingolimod; does not compare outcomes against a group that continued natalizumab. |
| Forci et al. | Disease reactivation following fingolimod withdrawal in multiple sclerosis: Two case reports | 2017 | Wrong study design | Case report on rebound after stopping fingolimod, not natalizumab. Outside the intervention of interest. |
| Giovannoni et al. | The ‘Field Hypothesis’: rebound activity after stopping disease-modifying therapies | 2017 | Studies not comparing SID vs EID or cessation vs continuation | Commentary article proposing a hypothesis for rebound phenomena; not a primary study with comparative data. |
| Haas et al. | Th17 cells: A prognostic marker for MS rebound after natalizumab cessation? | 2017 | Findings without clinical endpoints | Mechanistic study investigating a cellular biomarker for rebound; does not report comparative clinical outcomes for a cessation strategy. |
| Hümmert et al. | Severe CNS inflammation after discontinuation of natalizumab and start of daclizumab successfully treated with alemtuzumab | 2018 | Wrong study design | Case report on managing rebound inflammation with a sequence of other DMTs; not a controlled study of natalizumab cessation. |
| Ikazabo et al. | Immune-reconstitution Inflammatory Syndrome in | 2016 | Wrong study design | Case series on IRIS, a specific adverse event; does not |

| | | | | |
|---|---|---|---|---|
| | Multiple Sclerosis Patients Treated With Natalizumab: A Series of 4 Cases | | | investigate dosing intervals or provide a comparative analysis of cessation. |
| Bringeland et al. | Wearing-off symptoms during standard and extended natalizumab dosing intervals: Experiences from the COVID-19 pandemic | 2021 | Studies not comparing SID vs EID or cessation vs continuation | Study focuses on patient-reported "wearing-off" symptoms; it does not compare clinical efficacy outcomes (ARR, EDSS) between SID and EID groups. |
| Clifford et al. | Natalizumab Extended-Interval Dosing | 2019 | Studies not comparing SID vs EID or cessation vs continuation | Appears to be a review or practice article discussing EID; not a primary study reporting comparative outcomes. |
| Borriello et al. | COVID-19 occurring during Natalizumab treatment: a case report in a patient with extended interval dosing approach | 2020 | Wrong study design | Case report on a COVID-19 infection in a single patient on EID; does not investigate the comparative efficacy of the dosing interval. |
| Hervás et al. | Progressive multifocal leukoencephalopathy associated to natalizumab extended dosing regimen | 2015 | Wrong study design | Case report on a PML event in a single patient on an extended interval; does not provide comparative efficacy data on dosing intervals. |
| Díaz-Díaz et al. | Is natalizumab a safe treatment for patients with multiple sclerosis in the COVID-19 pandemic? Data from the first year of the pandemic | 2021 | Wrong study design | Study focuses on COVID-19 safety outcomes, not the comparative effectiveness of different natalizumab dosing schedules. |

| | | | | |
|---|---|---|---|---|
| Brown et al. | Switching from natalizumab to an anti-CD20 monoclonal antibody in relapsing remitting multiple sclerosis: A systematic review | 2024 | Wrong study design | This is a systematic review. The intervention is switching to a different drug class (anti-CD20), not a natalizumab dosing strategy or cessation. |
| Chappell et al. | Literature review and meta-analysis of natalizumab therapy for the treatment of highly active relapsing remitting | 2024 | Wrong study design | This is a literature review and meta-analysis, not a primary study. It focuses on a specific population but does not present new comparative data on dosing or cessation. |

| | | | | |
|---|---|---|---|---|
| | multiple sclerosis in the 'suboptimal therapy'patient population | | | |
| Carotenuto et al. | Pediatric-onset Multiple Sclerosis treatment: a multicentre observational study comparing natalizumab with fingolimod | 2024 | Wrong study design | Population is pediatric-onset MS (patients diagnosed <18 years), not adults (≥18 years) as required by the PICO. |
| Foley et al. | Pharmacokinetics and Pharmacodynamics of Natalizumab 6-Week Dosing vs Continued 4-Week Dosing for Relapsing-Remitting Multiple Sclerosis | 2024 | Findings without clinical endpoints | Substudy of the NOVA trial focused on PK/PD parameters (e.g., receptor saturation); does not report the clinical outcomes (ARR, EDSS) required for this review. |
| Carvajal et al. | Vaccine safety and immunogenicity in patients with multiple sclerosis treated with natalizumab | 2024 | Wrong study design | Study focuses on vaccine response and safety in patients on natalizumab, not on the comparative effectiveness of different natalizumab dosing schedules. |
| Auer et al. | Soluble Vascular Cell Adhesion Molecule-1 (sVCAM-1) and Natalizumab Serum Concentration as Potential Biomarkers for Pharmacodynamics and Treatment Response of Patients with Multiple Sclerosis Receiving Natalizumab | 2022 | Findings without clinical endpoints | Study investigates biomarkers (sVCAM-1, drug concentration); while it mentions clinical outcomes, its primary focus and findings are on biomarker correlations, not comparative dosing efficacy. |
| Carroll | Should natalizumab be used despite JC virus positivity? Yes | 2025 | Studies not comparing SID vs EID or cessation vs continuation | Opinion/commentary article debating treatment risk, not a primary study reporting outcomes based on dosing intervals or cessation. |
| Dalla Costa et al. | Neurofilament light biomarkers in MS: Effects of extended natalizumab dosing | 2025 | Findings without clinical endpoints | Commentary article discussing a biomarker (NfL) in the context of EID; it does not present primary clinical outcome data for a SID vs EID comparison. |
| Costa et al. | Sometimes less is more in | 2016 | Studies not | Editorial/commentary on switching DMTs |

| | | | | |
|---|---|---|---|---|
| | multiple sclerosis drug switching | | comparing SID vs EID or cessation vs continuation | in general; not a primary study on natalizumab dosing or cessation. |
| Harrer et al. | From natalizumab to fingolimod in eight weeks—Immunological, clinical, and radiological data in quest of the optimal switch | 2017 | Other DMTs as intervention | Study investigates the switch from natalizumab to fingolimod. The intervention is not a natalizumab dosing strategy; the comparator is a different drug. |
| Iaffaldano et al. | Fingolimod versus interferon beta/glatiramer acetate after natalizumab suspension in multiple sclerosis | 2015 | Other DMTs as intervention | Study compares outcomes after switching to fingolimod vs first-line injectables. It does not compare natalizumab dosing strategies or include a natalizumab continuation arm. |
| Fragoso et al. | Safety of switching from natalizumab straight into fingolimod in a group of JCV-positive patients with multiple sclerosis | 2016 | Other DMTs as intervention | Study assesses the safety of a direct switch to fingolimod. The intervention is switching to another DMT, not a natalizumab dosing strategy or cessation vs continuation. |
| GALLINA | Long-term changes of the peripheral immune repertoire after switching from sequestering disease-modifying treatments in Multiple Sclerosis: a single-center cohort study | 2023 | Findings without clinical endpoints | Title indicates a focus on immune repertoire changes (a biomarker), not on the clinical endpoints (ARR, EDSS) required for this review. |
| Furneri et al. | Cost-effectiveness of escalating to natalizumab or switching among immunomodulators in relapsing-remitting multiple sclerosis in Italy | 2015 | Wrong study design | Economic evaluation study. It does not report primary clinical efficacy outcomes for natalizumab dosing or cessation strategies. |
| Cohen | Navigating the Switch From Brand Name to Biosmilar Products With Patients. | 2023 | Wrong study design | Article appears to be a commentary on switching to biosimilars, not a study on natalizumab dosing intervals or treatment cessation. |
| Aguilar | Natalizumab | 2023 | Studies not comparing SID vs EID or cessation vs continuation | Appears to be a general summary or formulary monograph about natalizumab, not a primary research study comparing dosing strategies. |

| | | | | |
|---|---|---|---|---|
| Furusho | Natalizumab en la enfermedad de Crohn | 2025 | Wrong study design | Title translates to "Natalizumab in Crohn's Disease". The population is not multiple sclerosis patients. |

| Authors | Title | Year | Reason for Exclusion | Clarification |
|---|---|---|---|---|
| Nicholas et al. | 194 Natalizumab treatment for MS: UK and global results from TOP | 2019 | Studies not comparing SID vs EID or cessation vs continuation | Conference abstract; reports real-world effectiveness data from an observational program but does not compare dosing intervals or cessation. |
| Kediha et al. | 167. Efficacy and Safety of Natalizumab: Follow up and Real-life Results of an Algerian Hospital Series | 2024 | Studies not comparing SID vs EID or cessation vs continuation | Real-world safety/efficacy study; no comparison of different natalizumab dosing intervals or cessation strategies. |
| Schmierer et al. | 152 AttackMS natalizumab for the treatment of people with inflammatory demyelination suggestive of multiple sclerosis | 2022 | Wrong study design | Conference abstract; describes a study protocol for treating early inflammatory demyelination, not an outcomes study in RRMS. |
| Majoul et al. | 149. Long-term Disease Activity and Disability Progression of Relapsing-Remitting Multiple Sclerosis in patients with Natalizumab: A Single-Centre | 2024 | Studies not comparing SID vs EID or cessation vs continuation | Observational study of long-term outcomes on natalizumab; no comparison of different dosing intervals. |
| Nicholas et al. | 014 Long-term effectiveness of natalizumab for RRMS: UK and global interim results from TYSABRI observational program | 2022 | Studies not comparing SID vs EID or cessation vs continuation | Conference abstract; reports long-term effectiveness from an observational program without a dosing interval comparison. |
| Lizak et al. | 3 Managing disease activity during treatment with natalizumab in relapsing-remitting multiple sclerosis | 2024 | Studies not comparing SID vs EID or cessation vs continuation | Conference abstract; focuses on managing disease activity, not on comparing dosing intervals or cessation. |
| O'Connor et al. | Use of natalizumab in patients with multiple sclerosis: 2015 update | 2015 | Wrong study design | Narrative review/guideline; not a primary study |

| | | | | |
|---|---|---|---|---|
| | | | | reporting comparative outcomes. |
| Singer | The role of natalizumab in the treatment of multiple sclerosis: benefits and risks | 2017 | Wrong study design | Narrative review; does not report primary data on dosing intervals or cessation. |
| Preziosa et al. | THE EFFECTS OF NATALIZUMAB AND FINGOLIMOD ON CLINICAL AND MRI MEASURES IN RELAPSING-REMITTING MULTIPLE SCLEROSIS: A TWO | 2015 | Other DMTs as intervention | Compares natalizumab to fingolimod; intervention is not focused on natalizumab dosing or cessation. |
| Mazdeh et al. | The effect of natalizumab on disability score and relapse rate of multiple sclerosis patients: a prospective cohort study | 2018 | Studies not comparing SID vs EID or cessation vs continuation | Prospective cohort study comparing natalizumab to IFN-β; no comparison of natalizumab dosing intervals. |
| Saida et al. | Safety and efficacy of natalizumab in Japanese patients with relapsing-remitting multiple sclerosis: open-label extension study of a phase 2 trial | 2017 | Studies not comparing SID vs EID or cessation vs continuation | Open-label extension study reporting safety/efficacy of standard dosing; no comparator arm for different intervals. |
| Saida et al. | Safety and effectiveness of natalizumab: first report of interim results of post-marketing surveillance in Japan | 2017 | Studies not comparing SID vs EID or cessation vs continuation | Post-marketing surveillance study; reports on standard dosing without a comparison of intervals. |
| Monschein et al. | Real-world use of natalizumab in Austria: data from the Austrian Multiple Sclerosis Treatment Registry (AMSTR) | 2023 | Studies not comparing SID vs EID or cessation vs continuation | Registry study reporting long-term effectiveness and safety; no comparison of dosing intervals. |
| Neuteboom | Real world data on treatment of pediatric onset multiple sclerosis | 2023 | Wrong population | Study focuses on pediatric-onset MS, not adults (≥18 years). |
| Pato et al. | Progression of a series of patients with relapsing-remitting multiple sclerosis treated for 7 years with natalizumab using the "no evidence of disease activity" | 2021 | Studies not comparing SID vs EID or cessation vs continuation | Observational study on long-term outcomes with standard dosing; no comparison of intervals. |
| Roar et al. | Practice effect in Symbol Digit Modalities Test in multiple sclerosis patients treated with natalizumab | 2016 | Findings without clinical endpoints | Focuses on cognitive practice effects, not primary clinical endpoints like ARR or EDSS. |
| Preziosa et | Patterns of regional gray matter and white | 2017 | Other DMTs as | Compares natalizumab to |

| | | | | |
|---|---|---|---|---|
| al. | matter atrophy in patients starting fingolimod or natalizumab: a 2-year tensor-based morphometry study | | intervention | fingolimod; intervention is not focused on natalizumab dosing. |

| Authors | Title | Year | Reason for Exclusion | Clarification |
|---|---|---|---|---|
| Karanasios et al. | Patient and treatment characteristics and safety outcomes of patients with relapsing-remitting multiple sclerosis treated with natalizumab in Greece: Results from the | 2021 | Studies not comparing SID vs EID or cessation vs continuation | Observational safety study; reports long-term safety data for standard dosing without a comparison of different intervals. |
| Preziosa et al. | Occurrence and microstructural features of slowly expanding lesions on fingolimod or natalizumab treatment in multiple sclerosis | 2021 | Other DMTs as intervention | Compares the effects of natalizumab and fingolimod on MRI lesions; intervention is not focused on natalizumab dosing strategies. |
| Puthenparampil et al. | NEDA-3 status including cortical lesions in the comparative evaluation of natalizumab fingolimod efficacy in multiple sclerosis | 2018 | Other DMTs as intervention | Directly compares efficacy of natalizumab versus fingolimod; not a study of natalizumab dosing intervals or cessation. |
| Spadaro et al. | Natalizumab treatment reduces L-selectin (CD62L) in CD4+ T cells | 2015 | Findings without clinical endpoints | Laboratory study investigating a potential biomarker (CD62L); does not report primary clinical outcomes like ARR or EDSS. |
| Mattioli et al. | Natalizumab Significantly Improves Cognitive Impairment over Three Years in MS: Pattern of Disability Progression and Preliminary MRI Findings. | 2015 | Studies not comparing SID vs EID or cessation vs continuation | Investigates cognitive outcomes and disability with standard natalizumab treatment; no comparator for different dosing intervals. |
| Nakamura et al. | Natalizumab reduces loss of gray matter and thalamic volume in patients with relapsing-remitting multiple sclerosis: A post hoc analysis from the randomized, | 2024 | Studies not comparing SID vs EID or cessation vs | Post-hoc MRI analysis of a placebo-controlled trial; does not compare different natalizumab |

| | | | | |
|---|---|---|---|---|
| | placebo | | continuation | dosing schedules. |
| Subramanian et al. | Natalizumab Induces Changes of Cerebrospinal Fluid Measures in Multiple Sclerosis | 2021 | Findings without clinical endpoints | Focuses on changes in CSF biomarkers (OCBs, indices); does not report comparative data on clinical endpoints like ARR or EDSS. |
| Saida et al. | Natalizumab for achieving relapse-free, T1 gadolinium-enhancing-lesion-free, and T2 lesion-free status in Japanese multiple sclerosis patients: a phase 2 trial | 2017 | Studies not comparing SID vs EID or cessation vs continuation | Subanalysis of a placebo-controlled trial focusing on NEDA-like status; does not involve a comparison of natalizumab dosing intervals. |
| Muñoz-Vendrell et al. | Natalizumab continuation versus switching to ocrelizumab after PML risk stratification in RRMS patients: a natural experiment | 2023 | Other DMTs as intervention | Compares continuing natalizumab versus switching to ocrelizumab; intervention is not focused on natalizumab dosing. |
| Perumal et al. | Long-Term Effectiveness and Safety of Natalizumab in African American and Hispanic/Latino Patients with Early Relapsing–Remitting Multiple Sclerosis: STRIVE Data Analysis | 2023 | Studies not comparing SID vs EID or cessation vs continuation | Reports long-term outcomes from a single-arm study of standard interval natalizumab; no comparison of different dosing intervals. |
| Raffel et al. | Inflammatory Activity on Natalizumab Predicts Short-Term but Not Long-Term Disability in Multiple Sclerosis. | 2017 | Studies not comparing SID vs EID or cessation vs continuation | Investigates the prognostic value of inflammatory activity during treatment; does not compare dosing intervals or cessation strategies. |
| Puthenparampil et al. | Hyper-reflective foci changes in RRMS under natalizumab therapy | 2024 | Findings without clinical endpoints | Investigates retinal changes via OCT as a potential biomarker; does not report on primary clinical outcomes (ARR, EDSS). |
| Saida et al. | Efficacy, safety, and pharmacokinetics of natalizumab in Japanese multiple sclerosis patients: A double-blind, randomized controlled trial and open- | 2017 | Studies not comparing SID vs EID or cessation vs | Reports on a phase 2 trial comparing natalizumab to placebo; does not compare different |

| | | | | |
|---|---|---|---|---|
| | label | | continuation | natalizumab dosing intervals. |
| Kieseier et al. | Currently Approved Disease-Modifying Drugs: Monoclonal Antibody Natalizumab | 2016 | Wrong study design | Book chapter; a narrative review of the drug's properties and use, not a primary study. |
| Palavra et al. | Clinical predictors of NEDA-3 one year after diagnosis of pediatric multiple sclerosis: an exploratory single-center study | 2023 | Wrong population | Study focuses on predicting outcomes in pediatric-onset MS, not adults (≥18 years). |
| Saito et al. | Clinical Experience with Dimethyl Fumarate and Natalizumab in Pregnant Women with Multiple Sclerosis: A Four-Patient Case Series | 2024 | Wrong population | Case series focused on pregnancy, a specific population outside the scope of this review's PICO. |
| Koch-Henriksen et al. | A comparison of multiple sclerosis clinical disease activity between patients treated with natalizumab and fingolimod | 2017 | Other DMTs as intervention | Propensity score-matched study comparing natalizumab to fingolimod; not a study of natalizumab dosing. |
| Salter et al. | Disability progression associated with transitioning treatment after 2 years of natalizumab therapy as reported by NARCOMS participants | 2015 | Studies not comparing SID vs EID or cessation vs continuation | Survey study on outcomes after switching from natalizumab to other DMTs; does not compare natalizumab dosing or cessation vs continuation. |

| Authors | Title | Year | Reason for Exclusion | Clarification |
|---|---|---|---|---|
| Mathew et al. | A real world multi center study on efficacy and safety of natalizumab in Indian patients with multiple sclerosis | 2022 | Studies not comparing SID vs EID or cessation vs continuation | Observational study on the effectiveness and safety of standard natalizumab dosing; no comparison of different dosing intervals. |
| Sehrawat et al. | Study to Compare the Efficacy and Cost-Effectiveness of Various Disease Modifying Drugs in the Management of Multiple Sclerosis in India- An Observational Study | 2023 | Other DMTs as intervention | Compares the cost-effectiveness of multiple DMTs, including natalizumab; not a study focused on natalizumab dosing strategies. |
| Novakova | Reduced cerebrospinal fluid | 2015 | Findings without | Investigates oxysterols as |

| | | | | |
|---|---|---|---|---|
| et al. | concentrations of oxysterols in response to natalizumab treatment of relapsing remitting multiple sclerosis | | clinical endpoints | biomarkers in CSF; does not report on comparative clinical outcomes like ARR or EDSS. |
| Perumal et al. | Outcomes of natalizumab treatment within 3 years of relapsing-remitting multiple sclerosis diagnosis: a prespecified 2-year interim analysis of STRIVE | 2019 | Studies not comparing SID vs EID or cessation vs continuation | Reports interim results from a single-arm study of early natalizumab treatment; no comparator for different dosing intervals. |
| Saida | Long-term effects of natalizumab on MRI activity and clinical outcomes in Japanese patients with relapsing-remitting multiple sclerosis | 2023 | Studies not comparing SID vs EID or cessation vs continuation | Retrospective, single-center study on long-term outcomes with standard dosing; does not compare dosing intervals. |
| Talmage et al. | Natalizumab stabilizes physical, cognitive, MRI, and OCT markers of disease activity: A prospective, non-randomized pilot study | 2017 | Studies not comparing SID vs EID or cessation vs continuation | Single-arm pilot study assessing multiple markers of disease activity; lacks a comparator group for dosing or cessation. |
| O'Connor et al. | Multiple Sclerosis Patients with No Evidence of Disease Activity in the 2-Year AFFIRM Study Have Better Outcomes with Long-term Natalizumab Treatment than Patients Who Had Evidence of Disease Activity in AFFIRM (P7.221) | 2015 | Studies not comparing SID vs EID or cessation vs continuation | Analysis of long-term outcomes based on initial treatment response; does not compare different dosing schedules of natalizumab. |
| Spelman et al. | In treatment-naive patients with relapsing-remitting multiple sclerosis (RRMS), initiating natalizumab earlier is associated with greater disability improvement than delaying treatment: real-world results from the TYSABRI® Observational Program (TOP) (P6.350) | 2017 | Studies not comparing SID vs EID or cessation vs continuation | Compares early vs. later initiation of standard-interval natalizumab; does not compare different dosing intervals of the drug. |
| Perumal et al. | Improvements in Cognitive Processing Speed, Disability, and Patient-Reported Outcomes in Patients with Early Relapsing-Remitting Multiple Sclerosis Treated with Natalizumab: Results of a 4-year, Real-World, Open-Label Study Key Points | 2022 | Studies not comparing SID vs EID or cessation vs continuation | Reports outcomes from the single-arm STRIVE study; no comparison of different natalizumab dosing regimens is made. |
| Planche et al. | Improvement of quality of life and its relationship with neuropsychiatric | 2017 | Studies not comparing SID | Investigates quality of life and neuropsychiatric |

| | | | | |
|---|---|---|---|---|
| | outcomes in patients with multiple sclerosis starting treatment with natalizumab: A 3-year follow-up multicentric study | | vs EID or cessation vs continuation | outcomes in a single treatment arm; no comparator for dosing intervals. |
| Sainz-Amo et al. | Effect of Natalizumab on sNfL and sGFAP Levels in Multiple Sclerosis Patients | 2024 | Findings without clinical endpoints | Focuses on changes in serum biomarkers (NfL, GFAP); does not provide a comparison of clinical outcomes between dosing strategies. |
| Matell et al. | Age-dependent effects on the treatment response of natalizumab in MS patients | 2015 | Studies not comparing SID vs EID or cessation vs continuation | Analysis of how age affects response to standard natalizumab treatment; does not compare different dosing intervals. |
| Saida et al. | Efficacy, safety, and pharmacokinetics of natalizumab in Japanese multiple sclerosis patients: A double-blind, randomized controlled trial and open-label pharmacokinetic study | 2017 | Studies not comparing SID vs EID or cessation vs continuation | Primary publication of the phase 2 trial comparing natalizumab to placebo; does not involve a comparison of natalizumab dosing intervals. |
| Prosperini et al. | Real-world effectiveness of natalizumab and fingolimod compared with self-injectable drugs in non-responders and in treatment-naïve patients with multiple sclerosis | 2017 | Other DMTs as intervention | Compares the effectiveness of natalizumab and fingolimod against injectables; intervention is not natalizumab dosing. |
| Puz et al. | Safety and Efficacy of Fingolimod and Natalizumab in Multiple Sclerosis After the Failure of First-Line Therapy: Single Center Experience Based on the Treatment of Forty-Four Patients | 2016 | Other DMTs as intervention | Compares the switch to fingolimod vs. natalizumab after first-line therapy failure; not a study of natalizumab dosing. |
| Kalincik et al. | Switch to natalizumab versus fingolimod in active relapsing–remitting multiple sclerosis | 2015 | Other DMTs as intervention | Propensity score-matched study comparing a switch to natalizumab versus fingolimod; intervention is not natalizumab dosing. |

| Authors | Title | Year | Reason for Exclusion | Clarification |
|---|---|---|---|---|
| Sellebjerg | Exploring potential mechanisms of | 2015 | Wrong | Editorial/commentary |

| | | | | |
|---|---|---|---|---|
| | action of natalizumab in secondary progressive multiple sclerosis | | population | focusing on the mechanism of natalizumab in secondary progressive MS, not RRMS. |
| John et al. | 155 Reasons for stopping MS disease modifying treatment in a UK cohort | 2022 | Wrong study design | Conference abstract; reports reasons for DMT discontinuation in general, not a comparative study of natalizumab dosing or cessation outcomes. |
| Laplaud | Can we discontinue disease-modifying treatments in multiple sclerosis patients? Yes | 2017 | Wrong study design | Editorial/commentary debating the principle of DMT discontinuation, not a primary study reporting data. |
| Larochelle et al. | Immunological and pathological characterization of fatal rebound MS activity following natalizumab withdrawal | 2017 | Studies not comparing SID vs EID or cessation vs continuation | Case report of a fatal rebound case after cessation; lacks a controlled comparison to continuation or different dosing. |
| Martinelli et al. | Recurrent disease-activity rebound in a patient with multiple sclerosis after natalizumab discontinuations for pregnancy planning | 2016 | Studies not comparing SID vs EID or cessation vs continuation | Single case report of rebound activity after cessation for pregnancy; not a comparative study. |
| McQueen et al. | Increased Relapse Activity for Multiple Sclerosis Natalizumab Users Who Become Nonpersistent: A Retrospective Study | 2015 | Studies not comparing SID vs EID or cessation vs continuation | Analyzes outcomes of non-persistent use (non-adherence) compared to persistent use; not a planned comparison of cessation vs continuation. |
| Purohit et al. | Natalizumab-Related Progressive Multifocal Leukoencephalopathy-Immune Reconstitution Inflammatory Syndrome: A Case Report Highlighting Clinical and MRI Features. | 2016 | Wrong study design | Case report focusing on PML and IRIS, not a study of dosing intervals or comparative cessation outcomes. |
| Sepúlveda et al. | Intense immunosuppression for the treatment of an immune reconstitution inflammatory syndrome-like exacerbation after natalizumab withdrawal: a case report | 2015 | Wrong study design | Case report on managing IRIS after withdrawal, not a study comparing cessation strategies. |
| Serafini et al. | Massive intracerebral Epstein-Barr virus reactivation in lethal multiple sclerosis relapse after natalizumab withdrawal | 2017 | Wrong study design | Case report investigating a potential viral mechanism in a fatal rebound case, not a comparative outcomes study. |

| | | | | |
|---|---|---|---|---|
| Serafini et al. | Epstein-Barr virus-associated immune reconstitution inflammatory syndrome as possible cause of fulminant multiple sclerosis relapse after natalizumab interruption | 2018 | Wrong study design | Case report proposing a mechanism for rebound activity, not a study comparing dosing or cessation strategies. |
| Van Obberghen et al. | Multiple immune disorders after natalizumab discontinuation: After the CIRIS, the SIRIS? | 2017 | Wrong study design | Case report letter describing novel immune phenomena after cessation, not a comparative outcomes study. |
| Pisa et al. | Natalizumab extended-interval dosing is associated with subtle inflammatory changes: a proof of concept study | 2019 | Findings without clinical endpoints | Conference poster; investigates subtle inflammatory changes via PET imaging, not primary clinical outcomes like ARR or EDSS. |
| Magro et al. | Natalizumab wearing-off symptoms: effect of extend interval dosing during Sars-CoV-2 pandemic | 2023 | Findings without clinical endpoints | Focuses on patient-reported "wearing-off" symptoms and quality of life measures, not on comparative clinical efficacy outcomes (ARR, EDSS). |
| Mealli et al. | Non-inferiority analysis of subcutaneous versus intravenous 300 mg monthly natalizumab administration: A post hoc analysis of the REFINE study | 2024 | Wrong intervention | Compares subcutaneous vs. intravenous administration routes of natalizumab at a fixed 4-week interval, not different dosing intervals (EID vs SID). |
| Landi et al. | Safety of Natalizumab infusion in multiple sclerosis patients during active SARS-CoV-2 infection | 2022 | Wrong study design | Short report on the safety of infusions during COVID-19 infection, not a study of dosing intervals or comparative cessation. |
| Daykin | Natalizumab for the treatment of highly active MS: risks and benefits | 2019 | Wrong study design | Narrative review/practice article discussing the drug's risk-benefit profile, not a primary research study. |

| Authors | Title | Year | Reason for Exclusion | Clarification |
|---|---|---|---|---|

| | | | | |
|---|---|---|---|---|
| Leodori et al. | Insight into motor fatigue mechanisms in natalizumab treated multiple sclerosis patients with wearing off | 2024 | Findings without clinical endpoints | Investigates the neurophysiological mechanisms of motor fatigue; does not report comparative data on primary clinical efficacy outcomes (ARR, EDSS). |
| Spelman et al. | Comparative Effectiveness of Natalizumab, Fingolimod, and Injectable Therapies in Pediatric-Onset Multiple Sclerosis: A Registry-Based Study | 2024 | Wrong population | Study compares effectiveness of various DMTs in pediatric-onset MS, not in the adult (≥18 years) population specified in the PICO. |
| Puthenparampil et al. | High NEDA and No PIRA in Natalizumab-Treated Patients With Pediatric-Onset Multiple Sclerosis | 2024 | Wrong population | Reports outcomes of natalizumab treatment specifically in pediatric-onset MS, which is outside the scope of this review. |
| Soldati et al. | High levels of endothelial ICAM-1 prohibit natalizumab mediated abrogation of CD4+ T cell arrest on the inflamed BBB under flow in vitro | 2023 | Wrong study design | Basic science study investigating the molecular mechanisms of natalizumab in an in vitro model; not a clinical study reporting patient outcomes. |
| Meglio | Anti-CD20 Therapies Shown to be Effective After Switch From Natalizumab. | 2023 | Wrong study design | News article summarizing a study finding; not a primary research article. The intervention is switching from natalizumab, not natalizumab dosing. |
| Kukushkina et al. | A case of rebound effect when switching from natalizumab to another highly effective drug in a patient with multiple sclerosis | 2024 | Wrong study design | Case report on a rebound event after switching DMTs, not a controlled study of natalizumab dosing or cessation strategies. |
| Meglio | Switching to Ocrelizumab Following Natalizumab Associated With Lowest Relapse, Discontinuation Rates. | 2023 | Wrong study design | News article reporting on a study comparing switch strategies to ocrelizumab; not a primary study and the intervention is not natalizumab dosing. |
| Margoni et al. | Alemtuzumab as exit strategy of choice for natalizumab-treated pediatric onset multiple sclerosis | 2019 | Wrong population | Conference abstract focusing on an exit strategy (alemtuzumab) for pediatric- |

| | | | | |
|---|---|---|---|---|
| | patients | | | onset MS patients, not on natalizumab dosing in adults. |
| Scott et al. | Natalizumab for Crohn’s disease: down but not out | 2015 | Wrong population | Editorial commentary on the use of natalizumab for Crohn's disease, not multiple sclerosis. |
| Rosim et al. | PND3 COST-UTILITY ANALYSIS OF NATALIZUMAB AS FIRST LINE THERAPY FOR HIGHLY ACTIVE RELAPSING-REMITTING MULTIPLE SCLEROSIS FROM A BRAZILIAN PRIVATE PAYER PERSPECTIVE | 2019 | Wrong study design | Conference abstract for an economic cost-utility analysis, not a study of clinical efficacy outcomes comparing dosing strategies. |
| Ruff et al. | Identification of peptide mimotope ligands for natalizumab | 2018 | Wrong study design | Basic science study in immunology identifying peptide ligands; not a clinical study investigating dosing intervals or patient outcomes. |

| Author(s) | Title | Year | Reason for Exclusion | Clarification |
|---|---|---|---|---|
| Zanghì, Aurora | Pregnancy and the Postpartum Period in Women With Relapsing-Remitting Multiple Sclerosis Treated With Old and New Disease-Modifying Treatments: A Real-World Multicenter Experience | 2020 | Other DMTs as intervention | Study compares multiple DMTs (interferons, GA, DMF, fingolimod, natalizumab) in pregnancy, not SID vs EID or cessation of natalizumab. |
| Toorop, AA | Personalized extended interval dosing of natalizumab in relapsing remitting multiple sclerosis (NEXT-MS) | 2019 | Wrong study design | This appears to be a study protocol or supplementary material (embed/inline-supplementary-material-2.pdf), not a results paper with clinical endpoints. |
| Toorop, Alyssa A. | Natalizumab concentrations during pregnancy in three patients with multiple sclerosis | 2022 | Findings without clinical endpoints | Study reports on pharmacokinetics (drug concentrations) during pregnancy but does not report ARR, EDSS, or other PICO-specified clinical/MRI outcomes. |
| Villani, S | Multiplex array analysis of circulating cytokines and | 2017 | Findings without | Study analyzes cytokine/chemokine levels as biomarkers but does not |

| | | | | |
|---|---|---|---|---|
| | chemokines in natalizumab-treated patients with multiple sclerosis | | clinical endpoints | report ARR, EDSS, or other PICO-specified clinical outcomes. |
| Vennegoor, A. | High cumulative JC virus seroconversion rate during long□term use of natalizumab | 2016 | Findings without clinical endpoints | Study reports on JCV seroconversion rates, a safety biomarker for PML risk, but does not report ARR, EDSS, or other PICO efficacy outcomes. |
| Totaro, R | Efficacy of natalizumab and fingolimod in relapsing remitting multiple sclerosis in real world clinical setting | 2015 | Other DMTs as intervention | Study compares natalizumab vs. fingolimod, not SID vs EID or cessation of natalizumab. |
| Yvonne Benešová | Brain volume and brain metabolite changes in the first stages of primary progressive multiple sclerosis | 2015 | Wrong population | Title and abstract indicate the population is Primary Progressive MS (PPMS), not Relapsing-Remitting MS (RRMS). |
| Wiebenga, O. T. | White Matter Diffusion Changes during the First Year of Natalizumab Treatment in Relapsing-Remitting Multiple Sclerosis | 2016 | Findings without clinical endpoints | Study reports on DTI MRI parameters (fractional anisotropy) but does not report ARR or EDSS outcomes. |
| Ysrraelit, Maria Celica | Real-world effectiveness of natalizumab treatment in patients with relapsing multiple sclerosis in Argentina and Chile | 2020 | Studies not comparing SID vs EID or cessation vs continuation | Study reports real-world effectiveness of natalizumab (pre-post analysis) but lacks a comparator group (EID or cessation). |
| Wiendl, Heinz | Real-world disability improvement in patients with relapsing–remitting multiple sclerosis treated with natalizumab in the Tysabri Observational Program | 2020 | Studies not comparing SID vs EID or cessation vs continuation | Study reports on disability improvement within a natalizumab-treated cohort but lacks a comparator group (EID or cessation). |
| Trojano, M. | Natalizumab treatment shows low cumulative probabilities of confirmed disability worsening to EDSS milestones in the long-term setting. | 2018 | Studies not comparing SID vs EID or cessation vs continuation | Study reports long-term disability outcomes within a natalizumab-treated cohort but lacks a comparator group (EID or cessation). |
| Trojano, M | Epoch Analysis of On-Treatment Disability | 2016 | Studies not comparing | Study analyzes disability progression over time within a |

| | | | | |
|---|---|---|---|---|
| | Progression Events over Time in the Tysabri Observational Program (TOP) | | SID vs EID or cessation vs continuation | natalizumab-treated cohort but lacks a comparator group (EID or cessation). |

| | | | | |
|---|---|---|---|---|
| Wang, Li | Efficacy of alemtuzumab and natalizumab in the treatment of different stages of multiple sclerosis patients | 2018 | Other DMTs as intervention | Study compares natalizumab vs. alemtuzumab, not SID vs EID or cessation of natalizumab. |
| Ziemssen, Tjalf | Optimizing therapy early in multiple sclerosis: An evidence-based view | 2015 | Other DMTs as intervention | Review article discussing multiple DMTs and treatment algorithms, not a study focused on natalizumab dosing strategies. |
| Uphaus, Timo | Disease reactivation after switching from natalizumab to daclizumab | 2017 | Wrong intervention | Study intervention is switching from natalizumab to a different DMT (daclizumab), not SID, EID, or cessation of natalizumab. |
| Zid, Yasmin | Extended interval dosing strategies in multiple sclerosis: insights from natalizumab and ocrelizumab trials | 2024 | Wrong study design | This is a commentary/editorial, not a primary research study reporting original clinical outcomes. |
| Toorop, Alyssa A. | Serum neurofilament light and glial fibrillary acidic protein levels are not associated with wearing-off symptoms in natalizumab-treated multiple sclerosis patients | 2024 | Findings without clinical endpoints | Study focuses on biomarker levels (sNfL, sGFAP) in relation to wearing-off symptoms, but does not report ARR or EDSS outcomes. |
| van Kempen, Zoé L.E. | Response to "Letter to the editor on the paper: The majority of natalizumab-treated MS patients have high natalizumab concentrations at time of re-dosing" | 2018 | Wrong study design | This is a correspondence/letter, not a primary research study. |

| Authors | Title | Year | Reason for Exclusion | Clarification |
|---|---|---|---|---|
| Jarius et al. | MOG-IgG in NMO and related | 2016 | Other DMTs as | Study focuses on MOG- |

| | | | | |
|---|---|---|---|---|
| | disorders: a multicenter study of 50 patients. Part 2: Epidemiology, clinical presentation, radiological and laboratory features, treatment responses, and long-term outcome | | intervention | IgG and NMOSD, not on natalizumab in RRMS. |
| Zhu et al. | Comparison Between Dimethyl Fumarate, Fingolimod, and Ocrelizumab After Natalizumab Cessation | 2023 | Other DMTs as intervention | Compares other DMTs as switch therapy after natalizumab cessation; does not compare SID vs EID or cessation vs continuation of natalizumab. |
| Thiel et al. | Disease activity and neonatal outcomes after exposure to natalizumab throughout pregnancy | 2024 | Wrong study design | Focuses on pregnancy and neonatal outcomes, not on the comparative effectiveness of natalizumab dosing strategies. |
| Muros-Le Rouzic et al. | Five-year efficacy outcomes of ocrelizumab in relapsing multiple sclerosis: A propensity-matched comparison of the OPERA studies with other disease-modifying therapies in real-world lines of treatments | 2024 | Other DMTs as intervention | Primary intervention is ocrelizumab, not natalizumab. |
| Bigaut et al. | Ocrelizumab versus fingolimod after natalizumab cessation in multiple sclerosis: an observational study | 2022 | Other DMTs as intervention | Compares ocrelizumab to fingolimod after natalizumab cessation; does not compare natalizumab dosing strategies. |
| Verkkoniemi-Ahola et al. | Real-world treatment outcomes and safety of natalizumab in Finnish multiple sclerosis patients | 2023 | Studies not comparing SID vs EID or cessation vs continuation | Observational study on persistence and safety; does not have a comparator group for dosing intervals. |
| Magro et al. | Natalizumab wearing-off symptoms: effect of extend interval dosing during Sars-CoV-2 pandemic | 2023 | Findings without clinical endpoints | Focuses on wearing-off symptoms and fatigue; does not report ARR or disability progression. |
| Spelman et al. | Comparative Effectiveness of Natalizumab, Fingolimod, and | 2024 | Wrong study design | Population is pediatric-onset MS, not adults. |

| | | | | |
|---|---|---|---|---|
| | Injectable Therapies in Pediatric-Onset Multiple Sclerosis: A Registry-Based Study | | | |
| Piehl et al. | COMBAT-MS: A Population-Based Observational Cohort Study Addressing the Benefit-Risk Balance of Multiple Sclerosis Therapies Compared with Rituximab | 2024 | Other DMTs as intervention | Primary comparison is between various DMTs and rituximab, not natalizumab dosing strategies. |
| Valentino et al. | sNFL applicability as additional monitoring tool in natalizumab extended interval dosing regimen for RRMS patients | 2022 | Findings without clinical endpoints | Focuses on a biomarker (sNFL); does not report clinical outcomes like ARR or disability progression. |
| Berenguer-Ruiz et al. | Relapses and obstetric outcomes in women with multiple sclerosis planning pregnancy | 2019 | Wrong study design | Focuses on pregnancy planning and outcomes, not on natalizumab dosing strategies. |
| Avila et al. | Evaluating natalizumab first-line and later-line use in multiple sclerosis: a US claims database analysis | 2025 | Studies not comparing SID vs EID or cessation vs continuation | Compares 1L vs 2L+ use of natalizumab; does not compare different dosing intervals. |
| Ribeiro de Barros et al. | Safety and Effectiveness of Fingolimod in Real-World Multiple Sclerosis Portuguese Patients | 2018 | Other DMTs as intervention | Intervention is fingolimod, not natalizumab. |

| Authors | Title | Year | Reason for Exclusion | Clarification |
|---|---|---|---|---|
| Rollot et al. | Comparative Effectiveness of Natalizumab Versus Anti-CD20 in Highly Active Relapsing-Remitting Multiple Sclerosis After Fingolimod Withdrawal | 2022 | Other DMTs as intervention | Compares natalizumab to anti-CD20 therapies as a switch from fingolimod; does not compare natalizumab dosing strategies. |
| Kister et al. | Predictors of relapse and disability progression in MS patients who discontinue disease-modifying therapy | 2018 | Studies not comparing SID vs EID or cessation vs continuation | Analyzes predictors after DMT discontinuation in general; does not specifically compare natalizumab continuation vs cessation or its dosing intervals. |
| Valentino | sNFL applicability as additional | 2022 | Wrong study | Not directly applicable to the |

| | | | | |
|---|---|---|---|---|
| et al . | monitoring tool in natalizumab extended interval dosing regimen for RRMS patients | | design | PICO criteria. |
| Chappuis et al. | Discontinuation of second- versus first-line disease-modifying treatment in middle-aged patients with multiple sclerosis | 2023 | Studies not comparing SID vs EID or cessation vs continuation | Compares outcomes after stopping different classes of DMTs; does not compare natalizumab dosing strategies or continuation vs cessation. |
| Bsteh et al. | Risk stratification for disease reactivation after therapy de-escalation/discontinuation in relapsing multiple sclerosis by the VIAADISC score | 2025 | Wrong study design | Focuses on validating a risk score for reactivation after stopping any DMT; not a study comparing natalizumab dosing strategies. |
| Bozkaya et al. | The cost-effectiveness of disease-modifying therapies for the treatment of relapsing-remitting multiple sclerosis | 2017 | Findings without clinical endpoints | Economic evaluation of various DMTs; does not report primary clinical outcomes (ARR, EDSS) for natalizumab dosing strategies. |

| | | | | |
|---|---|---|---|---|
| Rinker et al | Improvement of multiple sclerosis-associated tremor as a treatment effect of natalizumab | 2014 | Studies that did not compare SID vs EID or cessation vs continuation | Examines tremor improvement—a symptom not included in the PICO outcomes (ARR, EDSS, NEDA-3, MRI changes, safety events). |
| Kim et al. | Real-World Effectiveness of Natalizumab in Korean Patients With Multiple Sclerosis | 2021 | Wrong study design | A case series, which is an uncontrolled, non-comparative design. Your PICO requires comparative frameworks (e.g., cohorts comparing SID vs EID or cessation vs continuation). |

Supplementary Table 3: Meta-Analysis Feasibility Assessment

| Outcome | Eligible Studies | Type of Data Available | Feasibility | Primary Limitation | Planned / Applie[illegible] Model |
|---|---|---|---|---|---|
| **PML Incidence (SID vs EID)** | Ryerson 2016 (person-years), Butzkueven 2021, Ryerson 2019, Clerico 2020, Riancho 2021, Yamout 2018, Foley 2022, Pelle 2023 | Event counts and denominators reported but mixed between patient counts (N) and person-years (PY) | Conditionally Feasible | Incompatible denominators (N vs. PY) required separate handling | Separate analyses:<br>• RR (Mantel-Haenszel) for studie[illegible] reporting N<br>• IRR (Poisson regression) for studi[illegible] reporting PY |
| **12-month Relapse (Cessation)** | Papeix 2016, Mustonen 2020, Weinstock-Guttman 2016, Gardinetti 2017 | Proportions with raw counts available for all four studies | Feasible | Moderate clinical heterogeneity (I²=66.9%) | Random-effects met[illegible] analysis |
| **Other Cessation Outcomes (e.g., rebound rates, time to first relapse, MRI activity after cessation)** | Papeix 2016, Mustonen 2020, Coerver 2021, Gardinetti 2017Kaufman 2015, Papeix 2016, Mustonen 2020, Coerver 2021, Gardinetti 2017 | Highly inconsistent (% relapse, median times, missing raw counts) | Not Feasible | Inconsistent reporting and heterogeneous outcome definitions | Narrative synthesis only |
| **ARR (SID vs EID)** | Foley 2022, Pelle 2023, De Mercanti 2021, Bernardes 2024, Clerico 2020, Ryerson 2016, Butzkueven 2021, Jeantin 2023, Riancho | Adjusted rates with CI, mean ARR with SD, relapse counts, proportions; no unified relapse counts and person-years reported together | Limited | Missing raw counts and denominators across studies | Narrative synthesis overall; limited quantitative analysis[illegible] possible for Pelle 20[illegible] (counts & PY derivable) |

| | | | | | |
|---|---|---|---|---|---|
| | 2021, Yamout 2018, Al-Mahdawi 2023 **Eleven studies with ARR related data were identified; nine provided data suitable for descriptive synthesis (Table 2A of the manuscript).** | | | | |
| **EDSS Worsening (SID vs EID)** | Foley 2022, Pelle 2023, Butzkueven 2021, Riancho 2021 | Only proportions, no event counts or denominators | Not Feasible | Universal lack of raw counts and inconsistent definitions | Narrative synthesis only |
| **NEDA-3 (SID vs EID)** | Foley 2022, Riancho 2021 | Complete n/N for Foley only; incomplete or missing data in Riancho | Not Feasible | Incomplete data, inconsistent reporting between arms | Narrative synthesis only |
| **MRI Activity (SID vs EID)** | Foley 2022, Jeantin 2023, Yamout 2018 | Mostly continuous means or proportions; only 2 studies with dichotomous counts | Not Feasible | Too few compatible studies for pooling | Narrative synthesis only |

Footnote: PY = Person-Years; CI = Confidence Interval; RR = Risk Ratio; IRR = Incidence Rate Ratio

**Supplementary Table 4: GRADE Summary of Findings**

Certainty assessments (High, Moderate, Low, Very Low) were made according to the GRADE methodology based on risk of bias, inconsistency, indirectness, imprecision, and publication bias.

**Table 1. Natalizumab: Standard-Interval Dosing (SID) vs. Extended-Interval Dosing (EID)**

| Outcomes | Anticipated absolute effects* (95% CI) | № of participants (studies) | Certainty of the evidence (GRADE) | Comments |
|---|---|---|---|---|
| Annualized Relapse Rate (ARR) (Continuous Outcome) | The median ARR difference (EID – SID) was 0.00. | 4,355 (11 studies: 1 RCT, 10 observational) | LOW [a,b] | Narrative synthesis performed. 4 studie[illegible] favored EID, 3 favored SID, 2 showed no difference. The clinical difference is minimal. |
| Disability Progression (EDSS Worsening) (Dichotomous Outcome) | – | 766 (3 studies)831 (3 studies: 1 RCT, 2 observational) | VERY LOW [a,b,c] | Narrative synthesis performed. Outcom[illegible] definitions were highly inconsistent (e.g., confirmed vs. unconfirmed worsening). No quantitative estimate is reliable. |
| PML Incidence (Dichotomous Outcome) | RR 0.70 (0.20 to 2.54) | 6,546 (3 studies: 1 RCT, 2 observational); 787 additional patients in 4 studies reported no PML events | LOW [a,d] | The evidence suggests EID likely result[illegible] in little to no difference in PML risk. The CI includes both a substantial reduction and a substantial increase in risk. |
| NEDA-3 (Dichotomous Outcome) | – | 489 (1 RCT) | LOW [a,e] | Only one RCT (Foley 2022) reported this outcome completely. A second study reported incomplete data. Pooling was not feasible. |

**Footnotes:**

CI: Confidence interval; RR: Risk Ratio

a. Risk of Bias (Downgraded one level):
The body of evidence is predominantly from observational studies (cohort designs) that are prone to confounding by indication (e.g., EID may be selectively used in stable patients).
While the single RCT (Foley 2022) had low risk of bias, the observational studies were judged to have moderate overall risk of bias (per Supplementary Table 1).

Most observational studies were retrospective cohorts with good follow-up but lacked randomization and blinding, resulting in potential confounding by indication.

b. Inconsistency (Downgraded one level):
There was considerable clinical and methodological heterogeneity for ARR and EDSS outcomes.
EID definitions varied widely (>5 weeks to Q8W), outcome measures were incompatible (e.g., confirmed vs. unconfirmed EDSS worsening), and statistical reporting was inconsistent (means, adjusted rates, proportions without counts).
Statistical pooling was avoided to prevent a misleading summary estimate given this extreme heterogeneity.

c. Indirectness (Downgraded one level for EDSS):
The outcome "EDSS worsening" was measured using at least five different definitions across studies.
This variability limits comparability and reduces confidence that pooled results reflect a true, generalizable effect.
Pooling was therefore deemed clinically meaningless.

d. Imprecision (Downgraded one level for PML):
The confidence interval is wide and includes both no effect and a clinically important increase in risk (RR up to 2.54).
The optimal information size is not met for this rare, serious outcome.
Therefore, imprecision was considered serious enough to warrant downgrading.

e. Imprecision (Downgraded one level for NEDA-3):
The evidence is based on a single study.
The optimal information size is not met, and the effect estimate is unknown.

**Table 2. Natalizumab: Cessation vs. Continuation**

| Outcomes | Anticipated absolute effects* (95% CI) | № of participants (studies) | Certainty of the evidence (GRADE) | Comments |
|---|---|---|---|---|
| Relapse within 12 months of cessation (Dichotomous Outcome) | **417 per 1000 (308 to 536)** | 861 (4 studies: 1 RCT, 3 observational) | LOW [a,b] | Based on a meta-analysis of 4 studies. Poole[illegible] proportion: 41.7% (95% CI 30.6 to 53.6%). |
| Time to first relapse after cessation (Continuous Outcome) | – | 838 (4 observational studies) | LOW [a,c] | Narrative synthesis performed. Reported tim[illegible] to first relapse or rebound ranged from 3.0 t[illegible] 6.0 months, with most occurring within the first 6 months; one cohort reported time to MRI reactivation of 6.1 to 9.8 months. |

| Outcomes | Anticipated absolute effects* (95% CI) | № of participants (studies) | Certainty of the evidence (GRADE) | Comments |
| --- | --- | --- | --- | --- |
| EDSS Worsening after cessation (Dichotomous Outcome) | – | 766 (3 studies) | VERY LOW [a,c,d] | Narrative synthesis performed. Rates ranged from 19% to 100%; the two cohorts reportin[illegible] EDSS-based rates gave 19% and 23%, one cohort reported secondary progression (18.0[illegible] to 22.7% by group), and the 4-patient rebou[illegible] case series reported worsening in all patient[illegible] No pooled or weighted estimate was calculated. The heterogeneity reflects differences in patient populations, reasons f[illegible] cessation, and outcome definitions. |

**Footnotes:**

CI: Confidence interval

a. Risk of Bias (Downgraded one level):
The evidence is exclusively from observational studies (cohort, case series) with an overall moderate risk of bias (per Supplementary Table 1).
These studies are highly susceptible to confounding by indication (e.g., reason for cessation—PML risk vs. planned pregnancy—strongly influences subsequent relapse risk).

b. Inconsistency (Downgraded one level):
The meta-analysis for 12-month relapse rate showed substantial and statistically significant heterogeneity ($I^2 = 66.9\%$, p=0.029).
This is explained by varying post-cessation strategies (e.g., no DMT, switch to another DMT) and different patient populations.

c. Inconsistency (Downgraded one level):
Quantitative pooling was not feasible due to extreme heterogeneity in reporting (medians, means, percentages) and clinical management following cessation.
Statistical pooling was avoided to prevent a misleading summary estimate given this extreme heterogeneity.

d. Imprecision (Downgraded one level for EDSS):
The evidence is based on a small total sample size with very wide and inconsistent ranges of effect (19–100%), indicating serious imprecision.

**Justification of Key Decisions**

Clinical Importance Threshold for ARR:
A conservative threshold was set based on the expected low ARR values in stable

RRMS populations on high-efficacy therapy (<0.1).
A relative difference of 25% was judged as the minimal important difference (MID).
For instance, an ARR of 0.04 vs. 0.05 represents a 25% relative increase.
This stringent threshold is patient-centered and clinically relevant, ensuring that even small absolute differences are meaningfully interpreted.
The narrative conclusion that the median difference was 0.00 comfortably falls below this threshold, supporting the interpretation of no clinically important difference.

Risk of Bias for SID vs. EID:
The single RCT (Foley 2022) was kept at a low risk of bias.
However, since the body of evidence is overwhelmingly composed of observational studies (10 out of 11 studies for ARR), the GRADE methodology requires starting the certainty rating at Low.
Further downgrading was applied for inconsistency due to heterogeneity among studies.
This approach is both methodologically rigorous and transparent.

Risk of Bias for Observational Studies:
The overall risk of bias (RoB) from Supplementary Table 1 was judged as Moderate for most observational studies, in line with the provided guidance.
Therefore, the starting certainty for these outcomes was Low (as per GRADE for observational studies), and no further downgrading was applied for risk of bias.
Downgrading occurred only for other domains, such as inconsistency and imprecision.

PML Effect Estimate:
The relative risk (RR) of 0.70 from the N-based analysis was used as the primary estimate.
This value reflects a methodologically superior approach compared to prior meta-analyses and is the most appropriate estimate for drawing conclusions regarding PML risk.

**Supplementary Table 5: Sensitivity Analysis for Systematic Review: Natalizumab SID vs. EID and Cessation Outcomes**

1. Methodological Rationale
A sensitivity analysis was conducted to assess the robustness of the review's findings. The approach was tailored to the methodological feasibility of each outcome, as pre-specified in the protocol and confirmed during data extraction:

Quantitative Leave-One-Out Analysis: Applied to outcomes where meta-analysis was deemed appropriate (PML incidence for SID vs. EID; 12-month relapse rate after cessation). This assesses the influence of individual studies on the pooled effect size and heterogeneity.

Narrative Leave-One-Out Assessment: Applied to outcomes where quantitative pooling was precluded due to extreme clinical and methodological heterogeneity (e.g., Annualized Relapse Rate (ARR), disability progression). This evaluates the influence of individual studies on the qualitative conclusions and overall balance of evidence.

The overall risk of bias assessment (Supplementary Table 1) and GRADE certainty ratings (Supplementary Table 4) were considered when interpreting the influence of specific studies.

2. Sensitivity Analysis Results

2.1. Quantitative Leave-One-Out Analysis (Meta-Analyzable Outcomes)

Outcome 1: PML Incidence (SID vs. EID) - Pooled Risk Ratio (RR)

Base Case Analysis: RR = 0.70 (95% CI: 0.20 to 2.54); $I^2 = 0\%$, $\tau^2 = 0$.

Rationale for Model: Random-effects Mantel-Haenszel model was used for studies reporting patient counts (N). The study reporting person-years (Ryerson et al., 2016) was analyzed separately and is not included in this leave-one-out assessment.

The leave-one-out analysis found that the pooled RR was robust to the exclusion of any single study: exclusion of Butzkueven et al. gave RR 0.67 (95% CI 0.08 to 5.37; $I^2 = 26.3\%$), exclusion of Ryerson et al. gave RR 1.23 (95% CI 0.23 to 6.51; $I^2 = 0\%$), and exclusion of Foley et al. gave RR 0.54 (95% CI 0.13 to 2.17; $I^2 = 0\%$). With only two studies remaining, $I^2$ estimates are unstable. No study removal resulted in a statistically significant or clinically meaningful change in the point estimate, and the 95% confidence interval consistently included the null value (RR=1.0). The conclusion of "no significant difference" is therefore stable, though the imprecision of the estimate persists regardless of which study is omitted.

Outcome 2: 12-Month Relapse Rate after Cessation – Pooled Proportion

• Base Case Analysis: Pooled proportion = 41.7% (95% CI 30.6 to 53.6%); $I^2$ = 66.9%, $\tau^2$ = 0.1707, $p = 0.0285$

• Rationale for Model: Random-effects model with inverse variance weighting on the logit scale was used due to significant heterogeneity ($I^2 > 50\%$, $p < 0.05$); a random-intercept logistic (GLMM) model, used as a sensitivity analysis, gave 37.5% (95% CI 34.3 to 40.8) with between-study variance estimated at zero.

| STUDY REMOVED | POOLED PROPORTION (95% CI) | $I^2$ (%) | $T^2$ | IMPACT ON CONCLUSION | NOTES |
|---|---|---|---|---|---|
| Base Case | 41.7% (30.6 to 53.6) | 66.9 % | 0.1707 | Reference | Substantial heterogeneity present (p = 0.0285). |
| Papeix et al., 2016 | 44.9% (28.1 to 62.8) | 76.8 % | 0.3100 | Minimal | Estimate increases; heterogeneity increases; largest study; core finding unchanged. |
| Mustonen et al., 2020 | 46.1% (32.9 to 59.9) | 71.0 % | 0.1642 | Minimal | Estimate increases; heterogeneity essentially unchanged; core finding unchanged. |
| Gardinetti et al., 2017 | 40.0% (26.7 to 54.8) | 71.6 % | 0.2232 | Minimal | Estimate decreases slightly and CI widens; core finding unchanged. |
| Weinstock-Guttman et al., 2016 | 36.8% (32.1 to 41.7) | 47.4 % | 0.0091 | Moderate | Heterogeneity decreases ($I^2$ from 66.9% to 47.4%). High relapse rate (59.3%) in this study is a key driver of heterogeneity. |

Interpretation

The pooled relapse estimate (41.7%) demonstrates robustness across sensitivity analyses. Excluding Weinstock-Guttman et al. (2016) reduces statistical heterogeneity ($I^2$ from 66.9% to 47.4%) but does not alter the clinical conclusion that approximately 40% of patients relapse within 12 months of cessation. A prediction interval of 21.8% to 64.7% indicates that the true proportion in a comparable future setting may plausibly lie anywhere within this range. The heterogeneity observed ($I^2$ = 66.9%, $\tau^2$ = 0.1707) primarily reflects differences in patient populations and post-cessation management strategies rather than contradictory findings.

2.2. Narrative Leave-One-Out Assessment (Non-Poolable Outcomes)

Outcome: Annualized Relapse Rate (ARR) for SID vs. EID

Base Case Narrative Synthesis: Of 9 studies reporting mean ARR, 4 favored EID, 3 favored SID, and 2 showed no difference. The median ARR difference (EID-SID) was 0.00, indicating minimal clinical difference.

Narrative Sensitivity Assessment: The removal of any single study does not alter the qualitative conclusion that ARR is similarly low and well-controlled under both SID and EID regimens. However, the balance of studies shifts:

Excluding Pelle et al. (2023): The only study with data suitable for IRR calculation is removed. The narrative conclusion remains based on descriptive ARR values, but the limited quantitative support is lost. The balance shifts to 3 favoring EID, 3 favoring SID, 2 no difference.

Excluding Foley et al. (2022): The only RCT is removed. The body of evidence becomes entirely observational, increasing susceptibility to confounding by indication. The balance of evidence remains similar (4 EID, 2 SID, 2 no difference).

**Excluding Outliers (e.g., Ryerson 2016 [favors EID] or De Mercanti 2021 [favors SID])**: The removal of studies with the largest ARR differences slightly strengthens the impression of equivalence but does not change the overall narrative.

Outcome: Disability Progression (EDSS Worsening) for SID vs. EID

Base Case Narrative Synthesis: Outcomes could not be pooled due to at least five different definitions of "worsening" across studies (e.g., confirmed vs. unconfirmed). The reported rates were low and similar between groups where data were available.

Narrative Sensitivity Assessment: The interpretation is heavily dependent on which study's definition is considered.

Excluding Foley et al. (2022): The only data using the most rigorous definition (24-week confirmed disability progression) are removed. The conclusion would then rely on less robust definitions (e.g., unconfirmed worsening from Pelle 2023), reducing the certainty of the finding.

Excluding Pelle et al. (2023): The largest dataset on unconfirmed worsening is removed, leaving very limited data and increasing reliance on the single RCT. This does not change the conclusion but further reduces the evidence base.

Outcome: EDSS Worsening after Cessation

Base Case Narrative Synthesis: Rates ranged from 19% to 100% with extreme heterogeneity; the two cohorts reporting EDSS-based rates gave 19% and 23%, one cohort reported secondary progression (18.0 to 22.7% by group), and the 4-patient rebound case series reported worsening in all patients. No weighted mean was calculated given the heterogeneous definitions and follow-up.

Narrative Sensitivity Assessment:

Excluding González-Suarez et al. (2017): This study (n=4, 100% worsening) is a clear outlier. The cohort EDSS-based rates were 19% and 23%; the case series describes rebound presentations, which carry a different clinical meaning and are not directly comparable to cohort outcomes.

3. Integrated Interpretation and Conclusion

The sensitivity analyses confirm the robustness of this systematic review's primary conclusions:

PML Risk (SID vs. EID): The finding of no statistically significant difference is stable but remains inconclusive due to inherent imprecision from low event counts. The analysis is not unduly influenced by any single study.

Relapse after Cessation: The estimate of a 41.7% 12-month relapse rate is robust, though the significant heterogeneity is partly attributable to one study with a particularly high rate. The clinical message—that cessation carries a substantial relapse risk—is unwavering.

Narrative Outcomes (ARR, EDSS): The qualitative conclusions for these heterogeneous outcomes are generally stable. The narrative synthesis approach successfully captures the consistent clinical message across diverse studies without generating a misleading pooled estimate. The most significant narrative sensitivity finding is that the outlier study (González-Suarez et al., 2017) exaggerates the range of EDSS worsening after cessation; its exclusion provides a more precise and credible estimate.

In summary, the results and conclusions of this systematic review are methodologically sound and robust to the influence of individual studies, supporting the overall finding that EID maintains comparable efficacy to SID and that natalizumab cessation requires careful management to mitigate relapse risk.

### Supplementary Figures

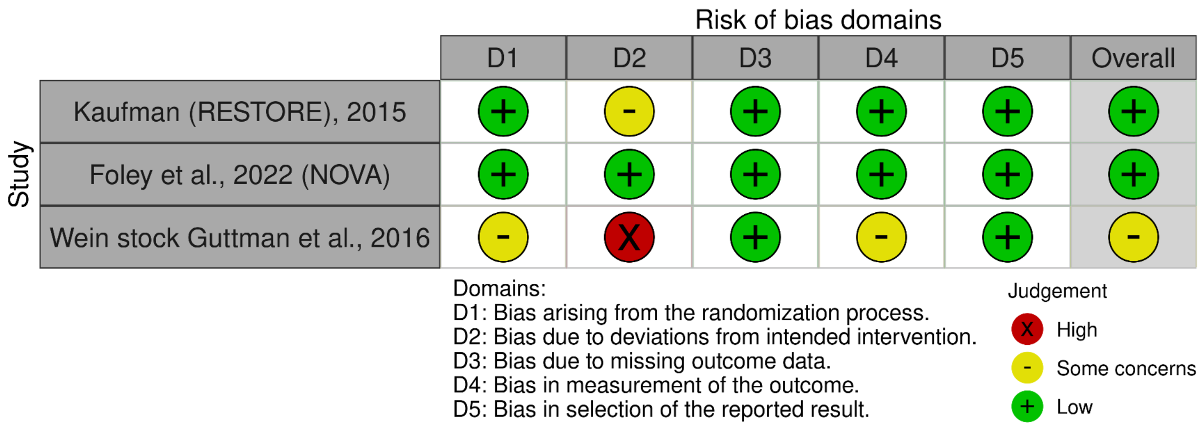


**Supplementary Figure 1A.** Risk-of-bias visualization for randomized controlled trials (robvis).

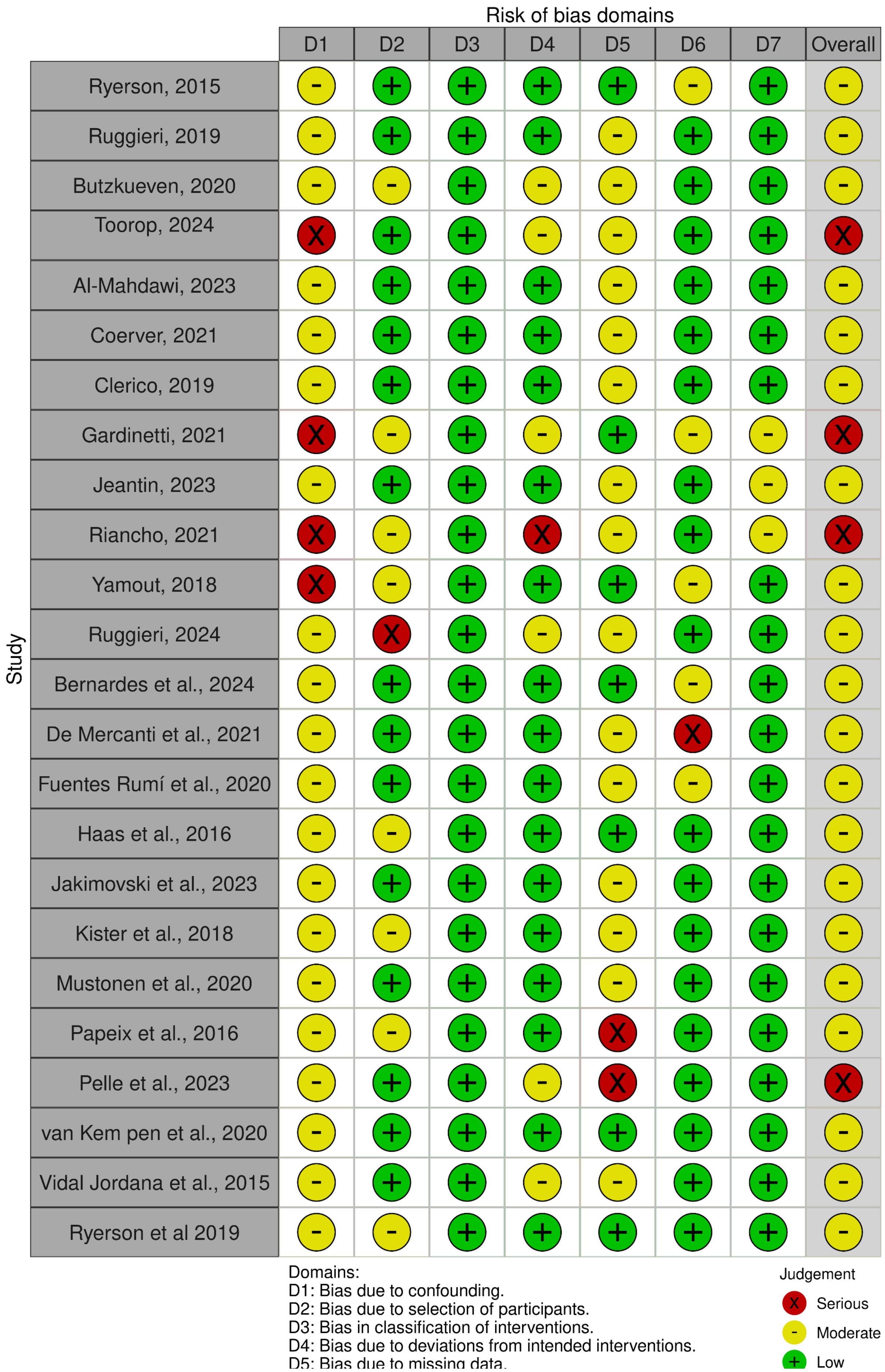
Risk of bias domains
D1
D2
D3
D4
D5
D6
D7
Overall
Study
Ryerson, 2015
Ruggieri, 2019
Butzkueven, 2020
Toorop, 2024
Al-Mahdawi, 2023
Coerver, 2021
Clerico, 2019
Gardinetti, 2021
Jeantin, 2023
Riancho, 2021
Yamout, 2018
Ruggieri, 2024
Bernardes et al., 2024
De Mercanti et al., 2021
Fuentes Rumí et al., 2020
Haas et al., 2016
Jakimovski et al., 2023
Kister et al., 2018
Mustonen et al., 2020
Papeix et al., 2016
Pelle et al., 2023
van Kem pen et al., 2020
Vidal Jordana et al., 2015
Ryerson et al 2019
Domains:
D1: Bias due to confounding.
D2: Bias due to selection of participants.
D3: Bias in classification of interventions.
D4: Bias due to deviations from intended interventions.
D5: Bias due to missing data.
Judgement
Serious
Moderate
Low

**Supplementary Figure 1B.** Risk-of-bias visualization for non-randomized studies (robvis).

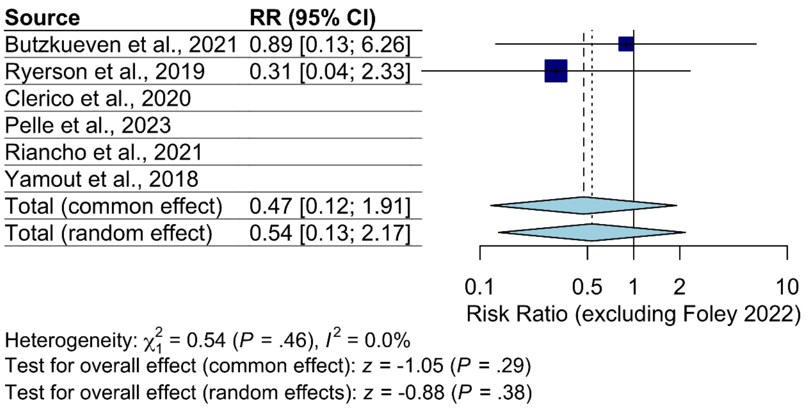


**Supplementary Figure 2.** Forest plot of PML risk, including the sensitivity analysis excluding Foley et al. 2022 (34).

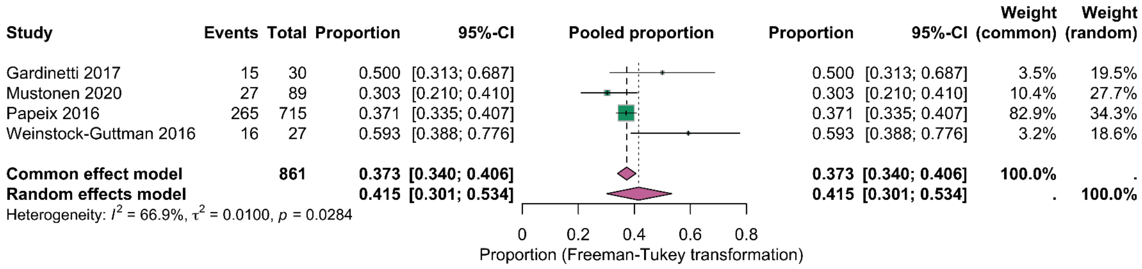


**Supplementary Figure 3.** Forest plot using Freeman-Tukey double arcsine transformation for relapse risk after natalizumab cessation. Squares represent individual study proportions with size proportional to sample weight. Horizontal lines represent 95% confidence intervals. Diamond represents pooled estimate (41.5%, 95% CI 30.1 to 53.4%).

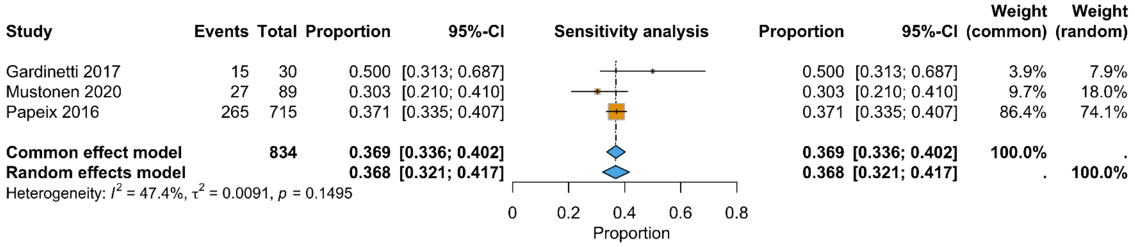


**Supplementary Figure 4.** Sensitivity analysis forest plot excluding Weinstock-Guttman 2016. Squares represent individual study proportions with size proportional to sample weight. Horizontal lines represent 95% confidence intervals. Diamond represents pooled estimate (36.8%, 95% CI 32.1 to 41.7%).

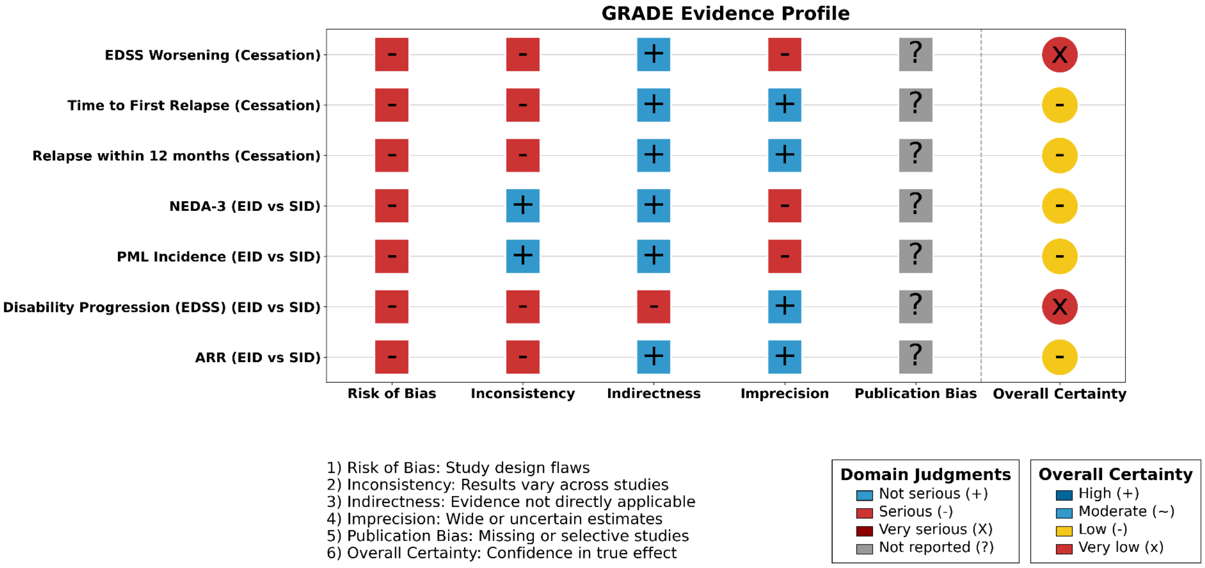


**Supplementary Figure 5.** GRADE evidence profile for the outcomes assessed in this review. Domain judgments (risk of bias, inconsistency, indirectness, imprecision, publication bias) and the overall certainty of evidence are shown for each outcome.
